# Multi-organ aging quantified from routine chest CT predicts chronic disease risk and mortality

**DOI:** 10.64898/2026.08.26.26361434

**Authors:** Junya Sato, Morteza Salehjahromi, Anas Zafar, Amgad Muneer, Xinyan Xu, Erjia Zhu, Natalie I. Vokes, Tina Cascone, Xiuning Le, Mehmet Altan, Eric E. Gardner, Ajay Sheshadri, Edwin J. Ostrin, Ameen A. Salahudeen, Tianhong Li, Miriam Merad, Aadel A. Chaudhuri, David E. Gerber, Fernando U. Kay, Myrna C.B. Godoy, Brett W. Carter, Girish S. Shroff, Lauren A. Byers, Caroline Chung, David Jaffray, David Rice, Zhongxing Liao, Joe Y. Chang, Ara A. Vaporciyan, Don L. Gibbons, Carol C. Wu, John V. Heymach, Jianjun Zhang, Jia Wu

## Abstract

Biological aging occurs heterogeneously across individuals and organs. However, current measures of biological age incompletely capture organ-specific differences in health and disease risk. Because chest CT visualizes multiple thoracic organs, it offers an opportunity to quantify structural aging across organ systems. Here, we developed MOSAIC-Age, a framework characterizing eight organ-specific aging clocks on chest CT. The clocks were developed and validated using 9,971 CT scans from CT-RATE and MIDRC, and subsequently locked and applied to two independent prospective cohorts with 35,293 participants from the National Lung Screening Trial and Genetic Epidemiology of COPD study. CT-derived biological age gaps (BAGs) were examined in relation to lifestyle and socioeconomic factors, prevalent comorbidities, incident chronic diseases, and all-cause and cause-specific mortality. Higher BAGs, indicating organs that appeared older on CT than expected for their chronological age, were broadly associated with adverse health characteristics, chronic disease burden, and increased mortality risk. Multiple disease outcomes were associated with aging across several organs, whereas in multivariable analyses including all eight organ-specific BAGs, the remaining associations were more organ specific. A greater number of markedly older-appearing organs and a faster pace of aging were each associated with higher mortality. Together, these findings demonstrate that routine chest CT captures both shared and organ-specific patterns of biological aging and establish CT-derived organ aging as a quantitative imaging biomarker for assessing multi-organ health and long-term disease risk.

## Introduction

Aging is a major contributor to chronic disease and mortality, reflecting the progressive loss of functional integrity across biological systems^1,2^. Biological aging varies substantially among individuals because it is shaped by genetic background^3^, lifestyle^4^, environmental exposures^5^, and sociodemographic factors^6^. Moreover, different organs within the same individual may age at different rates^7^. Although chronological age is widely used to assess aging-related risk, it does not necessarily capture the heterogeneity of biological aging across individuals and organs. Thus, characterizing organ-level biological aging may provide a more informative framework for disease risk stratification and personalized medicine than chronological age alone.

To move beyond chronological age, biological aging clocks have been developed from proteomic^8–11^, metabolomic^12^, and other omics approaches^13–15^. Although these studies have advanced the quantification of systemic aging, they characterize biological aging at the molecular level and do not directly quantify the structural manifestations of aging within specific organs. Imaging-based aging clocks offer a complementary, non-invasive means of quantifying age-related structural changes linked to organ function and disease^16^. However, to date, imaging-based aging studies have largely focused on brain MRI^17–20^, leaving structural aging in other organs less well characterized.

Imaging-based assessment of aging beyond the brain is especially important in the thorax, where age-related changes occur simultaneously across pulmonary, cardiovascular, and musculoskeletal systems. Important thoracic findings, including lung parenchymal abnormalities and vascular calcification, can be difficult to assess comprehensively with standard MRI protocols^21–23^. In contrast, CT provides high-resolution visualization of thoracic structures, and examination rates more than doubled between 2000 and 2016^24^. Given its widespread use for screening, diagnosis, and longitudinal follow-up^25,26^, routine chest CT offers a practical and scalable platform for opportunistic assessment of biological aging across thoracic organ systems; however, this potential has not been systematically investigated.

In this study, we developed MOSAIC-Age (Multi-Organ System Aging Clock), a deep learning-based thoracic aging clock trained using multicenter routine clinical chest CT scans (n=9,971) with no recorded abnormal findings. We evaluated its generalizability and clinical relevance in two prospectively collected cohorts with long-term follow-up, including 25,824 participants from the National Lung Screening Trial (NLST) and 9,469 participants from the Genetic Epidemiology of COPD (COPDGene) study. We then tested associations between the resulting organ-specific aging measures and chronic disease, mortality, and other health outcomes. Together, these analyses assess whether routine chest CT can support comprehensive, opportunistic assessment of multi-organ biological aging and aging-related health risk.

## Methods

### Study overview

The MOSAIC-Age (Multi-Organ System Aging Clock) framework was designed to capture organ-specific patterns of aging from CT images, with an end-to-end deep learning pipeline (**Fig. 1**): (1) each CT scan was segmented using a multi-organ segmentation model to generate anatomical masks for thoracic organs and structures, including lung, aorta, coronary artery, pulmonary artery, heart, bone, fat, and muscle. Bone, fat, and muscle were defined using predefined masks for the vertebrae; subcutaneous, torso, and intermuscular fat; and skeletal muscle, respectively; (2) following quality control, these masks were used to extract organ-specific three-dimensional CT crops; (3) deep learning models were trained to predict chronological age from each organ-specific crop; and (4) the organ-specific age difference between predicted age and chronological age was calculated to quantify whether an organ appeared older or younger on CT than expected for an individual’s age. We then evaluated associations between these organ-specific age differences and clinically relevant outcomes, including prevalent comorbidities, incident disease outcomes, and mortality.

**Fig. 1:**
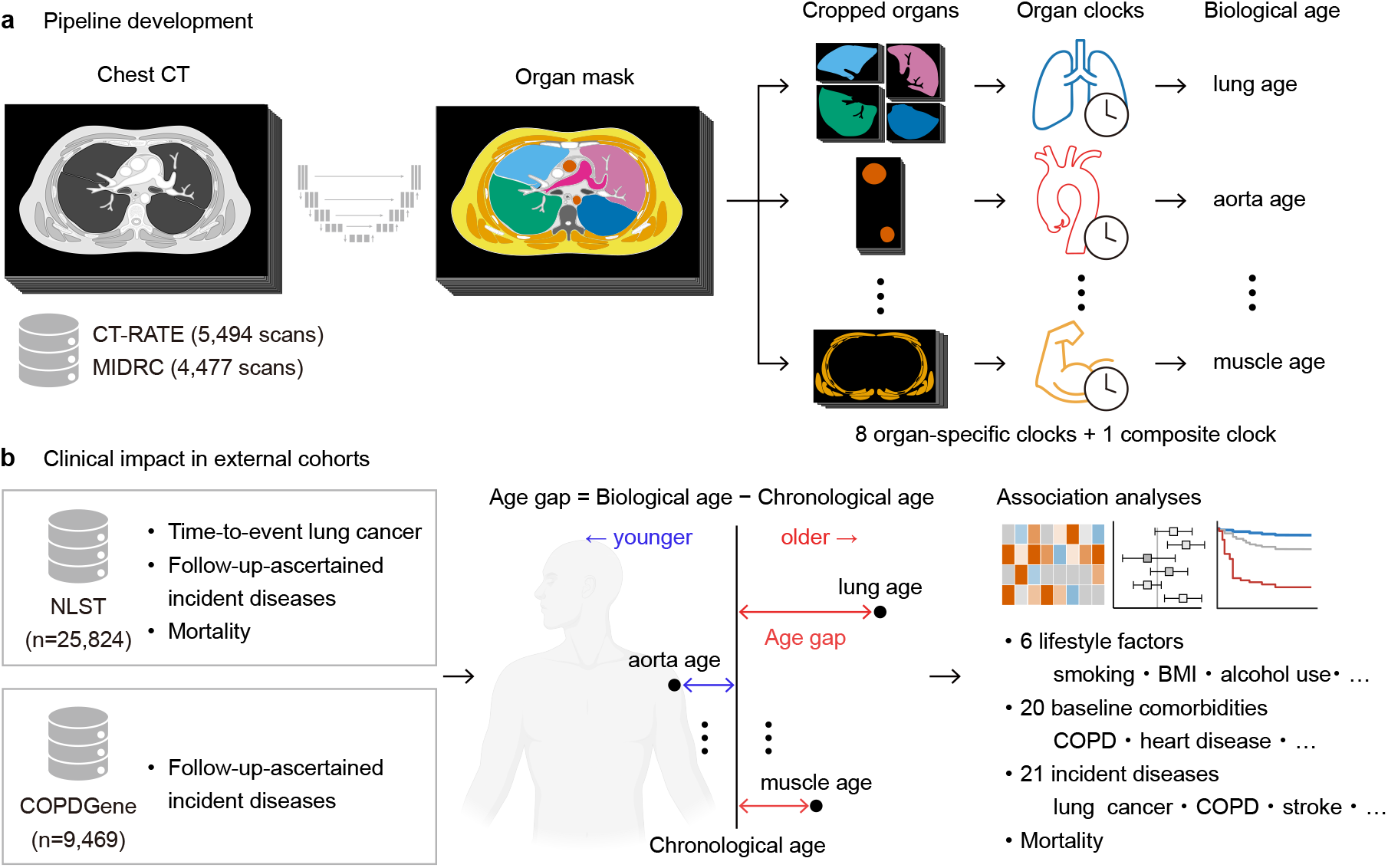
Study design for chest CT-based organ aging analysis. **a**, Overview of the end-to-end deep-learning pipeline for chest CT-based organ aging analysis. Chest CT images were first segmented using a pretrained multi-organ segmentation model, and organ-specific crops were extracted and used as inputs to the corresponding organ clock models. Organ clocks were developed using the CT-RATE and MIDRC datasets for eight anatomical targets: lung, aorta, coronary arteries, pulmonary artery, heart, bone, fat, and muscle, as well as a composite aging clock integrating the eight organ-specific predictions. **b**, Clinical evaluation of CT-derived organ aging. The trained pipeline was applied to the NLST and COPDGene cohorts to assess the clinical relevance of CT-derived organ aging. For each organ, a CT-derived biological age gap (BAG) was calculated from the predicted age and the participant’s chronological age. Associations between organ-specific BAGs and lifestyle factors, comorbidities, future disease risk, and mortality were then examined.

### Study cohorts

In this multicohort study, MOSAIC-Age was developed and validated using two independent chest CT datasets, the CT-RATE^27^ and a chest CT subset from the Medical Imaging and Data Resource Center (MIDRC)^28^. MOSAIC-Age was locked and clinically evaluated in two independent prospective cohorts, NLST and COPDGene. The study was approved by the Institutional Review Board of The University of Texas MD Anderson Cancer Center. Our retrospective secondary analysis of NLST and COPDGene was approved by their respective boards.

CT-RATE is a dataset of 50,188 chest CT volumes with radiology reports, derived from 25,692 examinations at Istanbul Medipol University Mega Hospital between May 2015 and January 2023^27^. Abnormality labels were extracted from the radiology reports. For organ-clock development, we used 5,494 CT images from 2,829 patients without recorded abnormal findings. This group provided a valuable baseline for exploring aging patterns in the absence of known clinical findings.

MIDRC is a multicenter medical imaging data commons established in 2020, containing de-identified imaging data, clinical information, and radiology reports from multiple institutions across the United States^28^. Among 8,855 chest CT images identified as of November 2025, 4,477 images from 4,083 patients without recorded disease information were included. These cases represent individuals with no reported abnormalities, offering an opportunity to study biological aging patterns in the absence of known clinical disease.

NLST is a multicenter lung cancer screening trial conducted at 33 sites in the United States between 2002 and 2007^29^. Between 2002 and 2004, the trial enrolled 53,454 high-risk smokers aged 55–74 years who had a smoking history of at least 30 pack-years and, if former smokers, had quit within the previous 15 years. Participants underwent three annual screening examinations, and approximately half were assigned to low-dose chest CT. For clinical evaluation of the proposed pipeline, 25,824 participants with an available baseline low-dose chest CT examination were included in the analysis. For longitudinal analyses, additional CT examinations from the second and third screening visits were collected when available.

COPDGene is a multicenter observational cohort designed to study the genetic, epidemiological and imaging determinants of chronic obstructive pulmonary disease^30^. Participants were current or former smokers aged 45–80 years with a smoking history of at least 10 pack-years. Between 2007 and July 2012, 10,371 participants were enrolled across 21 clinical centers in the United States. Chest CT was acquired for all participants according to the COPDGene imaging protocol, and de-identified images were transferred to the imaging core. For the proposed analysis, 9,469 participants with available CT images were included. In addition to CT imaging, COPDGene collected clinical and longitudinal follow-up data. Follow-up visits were conducted approximately every 5 years, during which newly developed diseases were recorded.

### Multi-organ system aging clock (MOSAIC-Age) framework

We developed and validated MOSAIC-Age, a deep learning framework designed to quantify organ-specific biological aging from routine chest CT scans. Separate organ clocks were developed after segmenting eight anatomical compartments with TotalSegmentator^31^, including the lung, aorta, coronary artery, pulmonary artery, heart, bone, fat, and muscle. Each organ clock was trained to learn normative age-related structural patterns and generate organ-specific age estimates from CT images. Organ-specific biological age gaps (BAGs) were subsequently calculated as the differences between predicted organ age and chronological age, providing a measure of accelerated or decelerated aging within each organ system. To summarize CT-based organ-specific aging as a global measure of biological aging, we additionally developed a composite multi-organ aging model using LightGBM, which took the eight organ-specific predicted ages as input features and generated a unified aging prediction. Detailed descriptions of model development, preprocessing, model architecture, and optimization are provided in **Supplementary Methods 1**.

The CT-RATE and MIDRC datasets were used for model development and validation. These datasets represent a geographically diverse population, including participants from multiple regions and clinical settings. Importantly, we restricted model training to individuals with no recorded abnormality labels, ensuring that the organ clocks captured age-related structural patterns rather than disease-specific signals. This resulted in 9,971 chest CT examinations from 6,912 individuals from two datasets. Additional details on the demographic and CT acquisition characteristics of each dataset are summarized in **Supplementary Table 1**. Because the two cohorts had different age distributions (mean [SD] age, 33.1 [9.7] years in CT-RATE and 60.1 [16.9] years in MIDRC), we combined them before splitting so that both training and testing reflected a broader age distribution. We then randomly split the combined dataset at the patient level into a 15% held-out test set and an 85% training set. Model tuning was performed within the training set using five-fold cross-validation. The stepwise ablation study used to determine the final model and the evaluation of model predictions across imaging modalities are shown in **Supplementary Methods 2** and **Supplementary Methods 3**, respectively.

### Calculation of CT-derived biological age gap (BAG)

For each organ-specific and composite aging clock, CT-derived BAGs were calculated using a regression-based correction to account for age-related prediction bias^9,32,33^. Specifically, for each clock, BAG was defined as the difference between a participant’s predicted age and the predicted age expected from the regression model for individuals of the same chronological age. This measure reflects whether a participant’s CT phenotype appears older or younger than expected for their age. Positive BAG values indicate older-appearing CT phenotypes relative to same-aged peers. BAGs were z-standardized within each organ-specific and composite measure before association analyses.

For participants with serial CT scans, BAG was calculated independently at each imaging visit. The rate of longitudinal change in BAG, referred to as BAG pace, was defined as the annualized change in BAG over time and was used to assess the relationship between aging pace and mortality risk.

To characterize extremes of biological aging, participants with organ-specific BAG values greater than 1.5 standard deviations above the cohort mean were classified as “extreme agers”, whereas those with BAG values more than 1.5 standard deviations below the mean were classified as “youthful agers”. These groups were subsequently evaluated for their associations with mortality outcomes.

### Participants’ baseline characteristics and comorbidities

Baseline demographic, lifestyle, socioeconomic, and clinical variables were obtained from the NLST and COPDGene datasets to evaluate associations with CT-derived BAGs. Variables included smoking pack-years, body mass index (BMI), alcohol use, marital status, educational attainment, and a binary occupational exposure indicator defined as any reported high-risk occupational exposure. Smoking pack-years and BMI were available in both datasets and analyzed in the combined cohort; the remaining variables were NLST-only and analyzed within the NLST cohort. The baseline comorbidities recorded in each dataset were also analyzed, including thoracic organ–related and systemic conditions for which at least 100 cases were available in the relevant dataset. The full list of comorbidities included in the analyses and their availability in NLST and COPDGene are provided in **Supplementary Table 2**. All variables corresponding to the baseline visit were associated with the baseline CT scan.

### Incident disease risk and mortality

Given differences in outcome availability between NLST and COPDGene datasets, primary analyses focused on the principal outcome supported by each dataset. In NLST, the availability of lung cancer diagnosis dates enabled time-to-event analysis of incident lung cancer. In COPDGene, genetic data permitted comparison of CT-derived lung aging with a previously developed lung-function polygenic risk score as a predictor of future risk of COPD^34^.

We then examined the relationship between organ-specific BAGs and a broader set of incident chronic diseases, including thoracic organ–related and systemic conditions for which at least 100 incident cases were available in the relevant dataset. For NLST, incident disease status was ascertained from follow-up reports and supplemented by underlying cause-of-death records for participants without the corresponding disease at baseline. For COPDGene, incident disease status was defined using disease-specific indicators collected at 5-year follow-up intervals. For outcomes with available baseline history, participants who had that disease at baseline were excluded from the disease-specific analysis. The full list of incident disease outcomes and their availability in NLST and COPDGene are provided in **Supplementary Table 3**.

Time-to-death and cause-of-death information were available in NLST and were used for the mortality analyses. All-cause mortality was the primary mortality endpoint. Cause-specific mortality was classified according to prespecified ICD-10 categories based on the underlying cause of death. Detailed definitions are provided in **Supplementary Table 4**.

For participants with serial CT examinations in NLST, the longitudinal change in BAG, referred to as BAG pace, was used to estimate the pace of biological aging. BAG pace was estimated using linear mixed-effects models fitted to BAG values from three annual CT scans and was defined as the annualized rate of change in BAG. Follow-up for BAG pace analyses began after the third annual CT scan used to estimate BAG pace, and participants who died before this scan were excluded.

### Statistical analysis

Associations between BAGs and baseline lifestyle and socioeconomic factors were assessed using linear regression, and associations between BAGs and baseline comorbidities were assessed using logistic regression. Cox proportional hazards regression was used for incident lung cancer and mortality outcomes with available event-time data. Other incident diseases were analyzed using logistic regression to model disease occurrence during follow-up. Models were adjusted for age, sex, and smoking pack-years, as applicable. When comparable variables or outcomes were available across both cohorts, analyses were pooled with additional adjustment for cohort; otherwise, analyses were performed within the relevant cohort. To evaluate the independent contribution of each organ-specific BAG rather than global aging processes shared across organs, models for incident disease and mortality were further adjusted for the remaining organ-specific BAGs.

For visualization of time-to-event associations, participants were classified into low, average and high BAG groups. Low BAG was defined as the lowest quartile of the BAG distribution, high BAG as the highest quartile, and average BAG as the middle 50%. Kaplan–Meier curves and cumulative incidence curves were generated for outcomes with event-time data stratified by BAG groups.

All statistical tests were two-sided. P values for the associations tested within each analysis were adjusted using the Benjamini–Hochberg method^35^. Adjusted P values are referred to as q values. Unless otherwise specified, statistical significance was defined as q < 0.05 after multiple-testing correction; P < 0.05 was used for analyses not subject to multiple-testing correction. For visualization of incident disease associations, absolute approximate Wald z statistics derived from the odds ratios (ORs) and 95% confidence intervals were normalized across BAGs with q < 0.05 within each outcome to sum to 1.

All statistical analyses were performed using Python (v3.12.11). Cox proportional hazards models were fitted using lifelines (v0.30.3). Linear and logistic regression models were fitted using statsmodels (v0.13.2). Pearson’s chi-square tests for independence were performed using scipy.stats (v1.13.1), and Benjamini–Hochberg false discovery rate correction was performed using statsmodels.stats.multitest. Additional libraries used for deep learning models are described in **Supplementary Method 4**.

## Results

### Robust age prediction performance of CT-based MOSAIC-Age

We developed the MOSAIC-Age framework using 9,971 chest CT scans from 6,912 participants in CT-RATE and MIDRC without apparent imaging abnormalities (**Supplementary Method 1** and **Extended Data Table 1**). In the held-out test set, the predicted age was closely correlated with the chronological age across organs (median Pearson r = 0.94; range, 0.81 to 0.97). Individual organ clocks predicted chronological age with a median mean absolute error (MAE) of 5.14 years across eight organs, whereas the composite clock achieved an MAE of 3.70 years (**Fig. 2a**). These MAEs were broadly comparable to those reported for prior MRI-based organ-specific aging clocks across multiple organs^36^.

**Fig. 2:**
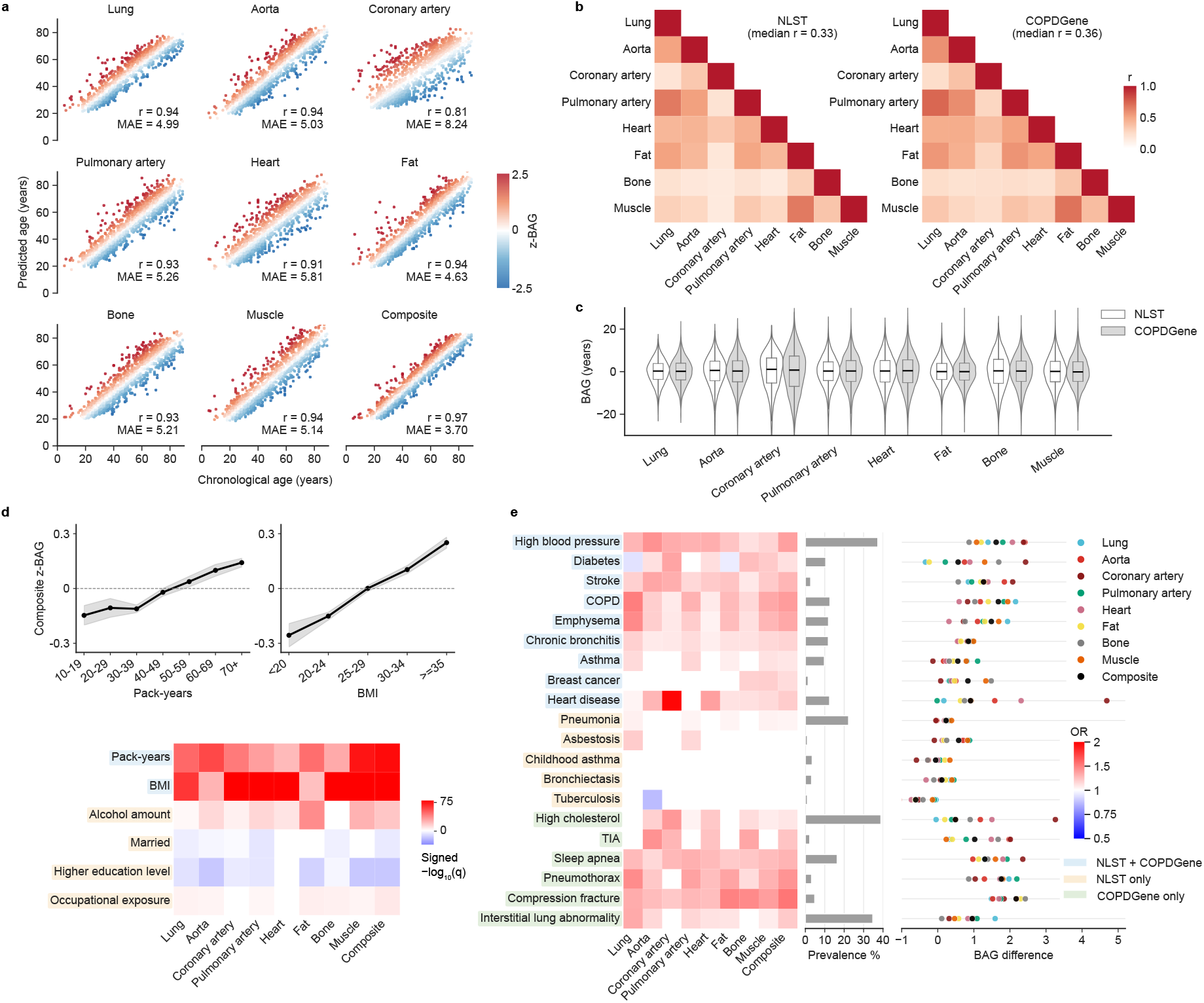
Performance of organ-specific aging clocks and associations with lifestyle factors and comorbidities. **a**, Scatter plots of predicted age versus chronological age for eight organ-specific aging clocks and the composite aging clock. The results are shown in the held-out (i.e. 15%) test dataset, with Pearson’s correlation coefficients and MAEs reported for each model. Point colors indicate the z-standardized BAG, with red and blue representing organs that appeared relatively older and younger than expected for chronological age, respectively. **b**, Pairwise correlations among organ-specific BAGs in NLST and COPDGene. **c**, Distributions of organ-specific BAGs, shown as violin plots with overlaid box plots. Boxes show IQR, center lines show medians, and whiskers extend to values within 1.5 × IQR. **d**, Associations between BAGs and lifestyle, socioeconomic, and environmental factors. Linear regression models were adjusted for age and sex. Line plots show associations of pack-years and BMI with the z-standardized composite BAG. Heatmap colors indicate signed −log_10_(q) values across organ-specific and composite BAGs; red and blue indicate positive and negative associations, respectively. Only associations with q < 0.05 are color coded. **e**, Associations between BAGs and prevalent comorbidities. Logistic regression models were adjusted for age, sex, and pack-years. Heatmap colors indicate odds ratios for organ-specific and composite BAGs. Bar plots show comorbidity prevalence, and dot plots show mean BAG differences between participants with and without each comorbidity. Background colors behind comorbidity labels indicate whether each comorbidity was available in both cohorts or in only one cohort. Only associations with q < 0.05 are color coded. BAG, biological age gap; BMI, body mass index; IQR, interquartile range; MAE, mean absolute error.

We next evaluated the robustness of MOSAIC-Age across multiple settings (**Supplementary Method 3**). The prediction performance was highly consistent between cross validation and the independent held-out test set for all eight organ-specific clocks as well as the composite model (**Extended Data Fig. 2a**). Age-prediction accuracy was comparable between male and female participants (median absolute difference in MAE, 0.35 years; **Extended Data Fig. 2b**). Finally, age estimates derived from diagnostic CT with contrast and the CT component of PET/CT showed moderate-to-strong agreement across organs (median Pearson’s r = 0.84; **Extended Data Fig. 2c**), supporting the robustness of MOSAIC-Age across commonly acquired clinical CT protocols.

### MOSAIC-Age reveals reproducible and organ-specific aging patterns linked to baseline health

MOSAIC-Age was locked and applied to baseline chest CT scans from two independent prospective multi-center cohorts with longitudinal follow-up: NLST (25,824 participants) and COPDGene (9,469 participants). The participants’ clinical characteristics are shown in **Extended Data Table 2**. CT-derived BAGs were calculated as age-adjusted residuals from regressions of CT-predicted age on chronological age, with positive values indicating organs that appeared older on CT than expected for individuals of the same chronological age. Representative CT images from participants with low and high BAGs are provided in **Supplementary Fig. 1**.

Pairwise correlations between organ-specific BAGs were generally weak in both cohorts (median Pearson r = 0.33 in NLST and 0.36 in COPDGene), except for higher correlations in the fat–muscle pair (NLST: r = 0.65; COPDGene: r = 0.68) and lung–pulmonary artery pair (NLST: r = 0.65; COPDGene: r = 0.73) (**Fig. 2b**). The generally weak correlations suggest partially shared but substantially organ-specific aging patterns, consistent with prior plasma proteomic organ-aging work^32^. Notably, the overall correlation structure in the heatmap as well as the distribution of organ-specific BAGs (**Fig. 2c**) were highly consistent across the two independent cohorts, supporting the robustness and reproducibility of CT-derived multi-organ aging patterns.

We next examined the associations between CT-derived BAGs and lifestyle, socioeconomic, and environmental factors. After multiple-testing correction, greater smoking pack-years, higher BMI, higher alcohol use, and reported occupational exposure were associated with higher composite BAGs, whereas being married and higher educational attainment were associated with lower composite BAGs (**Fig. 2d**). Among these factors, pack-years, BMI, and alcohol use were associated with the largest number of organ-specific BAGs (8/8, 8/8, and 7/8 organs, respectively), suggesting that lifestyle-related factors contribute broadly to CT-visible aging across multiple organs.

We then evaluated associations between BAGs and comorbidities that were present at the time of CT imaging. In logistic regression models adjusted for chronological age, sex, and pack-years, we tested 180 disease-BAG associations (20 comorbidities × 9 BAGs). After multiple-testing correction, 119 associations (66.1%) were significantly positive, indicating that higher CT-derived BAGs were broadly associated with higher odds of established chronic disease (**Fig. 2e**). Several associations followed expected organ–disease patterns, including lung BAG with prevalent COPD (OR per 1-s.d. higher BAG, 1.50; q = 2.4 × 10^−95^), coronary artery and heart BAGs with prevalent heart disease (coronary artery BAG: OR, 1.98; q = 9.9 × 10^−226^; heart BAG: OR, 1.37; q = 2.2 × 10^−72^), and bone BAG with prevalent compression fracture (OR, 1.46; q = 1.4 × 10^−12^). Associations were also observed outside the primary organ system of the disease, indicating that established chronic disease was accompanied by multi-organ patterns of CT-visible aging. Although these analyses were cross-sectional and do not establish causality, they demonstrate that MOSAIC-Age is able to capture clinically meaningful variation in baseline health and aging.

### MOSAIC-Age predicts incident chronic disease risk

We next assessed the prognostic value of baseline BAGs calculated by MOSAIC-Age for predicting future incident disease. In the NLST, where dates of lung cancer diagnosis enabled time-to-event analyses, six of the nine BAGs were associated with higher risk of incident lung cancer using Cox proportional hazards models adjusted for chronological age, sex, and pack-years (**Fig. 3a**). Among these, aorta BAG showed the largest observed association (HR per 1-s.d. higher BAG, 1.17; q = 1.9 × 10^−5^), while lung BAG was also associated (HR, 1.09; q = 0.013). Participants with high aorta BAG were more likely to develop lung cancer than those in the low or average aorta BAG groups (high versus low: HR, 1.60; P = 2.8 × 10^−7^; high versus average: HR, 1.29; P = 3.9 × 10^−4^; **Fig. 3b**). When all eight organ-specific BAGs were included in the same Cox model, only aorta BAG remained significant, whereas the other organ-specific BAGs no longer met the significance threshold (**Extended Data Fig. 3a**).

**Fig. 3:**
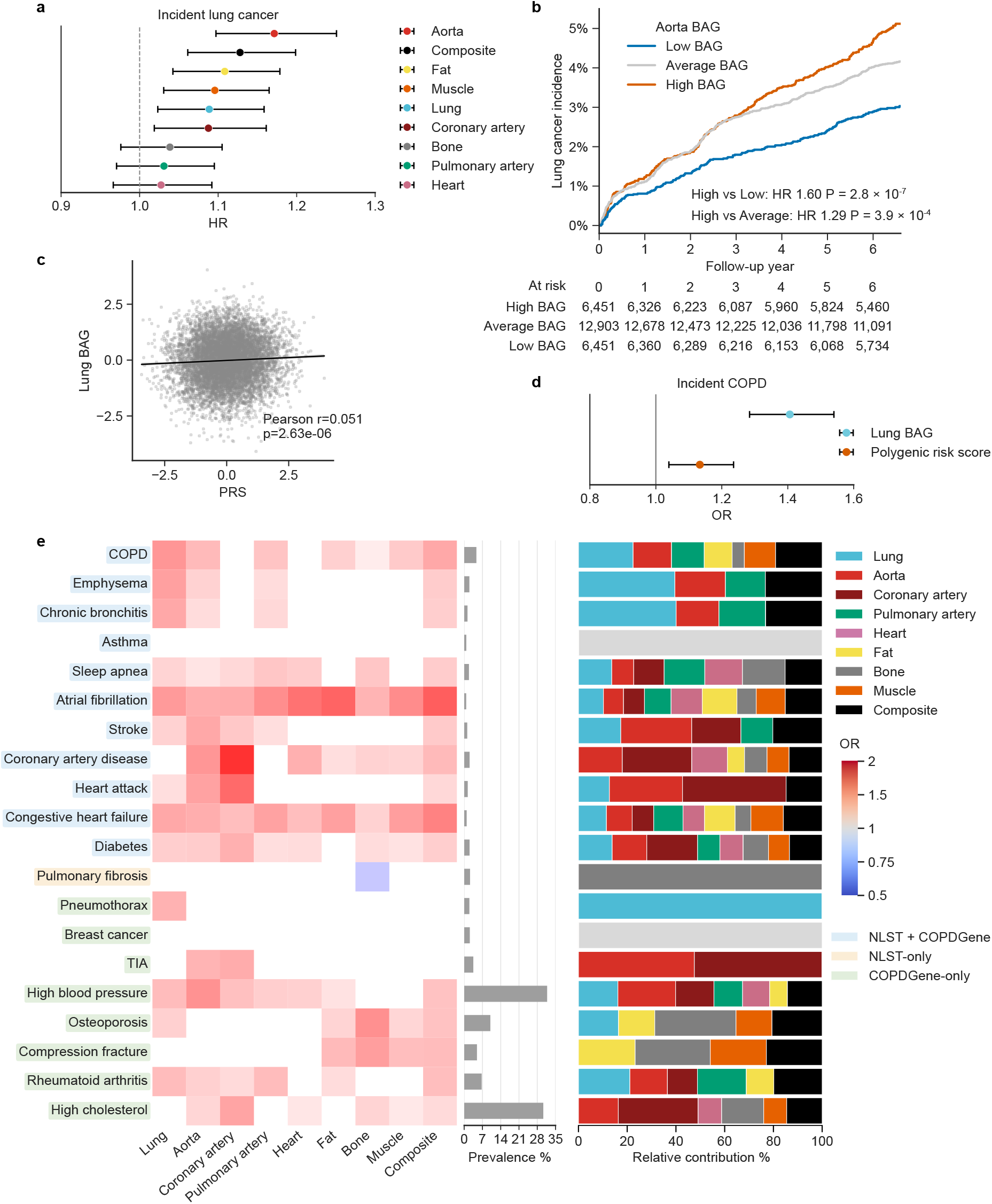
Associations between CT-derived BAGs and incident disease. **a**, Associations between CT-derived BAGs and incident lung cancer. **b**, Cumulative incidence of lung cancer stratified by aorta BAG. HRs for high versus low and high versus average BAG categories were estimated using Cox proportional hazards models adjusted for chronological age, sex, and smoking pack-years. **c**, Correlation between z-standardized lung BAG and lung function PRS, with Pearson r and P value shown. **d**, Associations of lung BAG and lung function PRS with incident COPD. **e**, Associations between BAGs and incident chronic diseases across 20 outcomes. In **a** and **d,** effect estimates are shown per 1-s.d. increase in the corresponding predictor. Cox proportional hazards regression was used for incident lung cancer in **a** and logistic regression for incident COPD in **d**; both models were adjusted for chronological age, sex, and pack-years. Horizontal lines indicate 95% confidence intervals. **e**, Logistic regression models tested associations between each organ-specific or composite BAG and incident disease, adjusting for chronological age, sex, and pack-years. Heatmap colors indicate odds ratios; only associations with q < 0.05 are color coded. Bar plots show the frequency of each incident disease outcome, and stacked bar plots show the normalized association magnitude of each significant BAG for each outcome, calculated by normalizing the absolute approximate Wald z statistics across BAGs with q < 0.05 so that they sum to 1. BAG, biological age gap; COPD, chronic obstructive pulmonary disease; PRS, polygenic risk score; TIA, transient ischemic attack.

To benchmark CT-derived lung aging against inherited genetic susceptibility, we compared lung BAG with a previously reported lung function polygenic risk score (PRS)^34^ in the COPDGene dataset. Lung BAG and the PRS were only weakly correlated (Pearson’s r = 0.051; **Fig. 3c**). After adjustment for chronological age, sex, and pack-years, lung BAG showed a stronger association with incident COPD than the PRS (OR per 1-s.d. increase, 1.41 versus 1.13; **Fig. 3d**). These findings suggest that lung BAG captures COPD risk information that was largely distinct from the genetic susceptibility.

We then extended the analysis to 20 chronic diseases across the nine BAGs in the NLST and COPDGene datasets. Of the 180 disease-BAG associations (20 incident diseases × 9 BAGs), 95 associations (52.8%) were significantly positive after multiple-testing correction (**Fig. 3e**), indicating that accelerated organ aging is broadly related to future chronic disease risk. For the disease risks examined, organ-specific BAGs showed larger effect sizes than the composite BAG. When all eight organ-specific BAGs were entered simultaneously into multivariable logistic regression models, the number of significant associations decreased and the remaining associations became more disease-specific (**Extended Data Fig. 3b**), suggesting that individual organ BAGs capture both shared and organ-specific components of biological aging. Together, these findings establish BAGs estimated by MOSAIC-Age as prospective biomarkers of chronic disease risk and demonstrate that organ-specific aging provides clinically informative signals beyond a single composite measure of biological age.

### MOSAIC-Age predicts all-cause and cause-specific mortality

We further examined whether CT-derived BAGs from MOSAIC-Age were associated with all-cause mortality. In Cox proportional hazards models adjusted for chronological age, sex, and pack-years, higher BAGs were broadly associated with increased mortality risk across all CT-derived BAG measures (**Fig. 4a**). Kaplan–Meier analysis demonstrated graded separation of all-cause survival across low, average, and high composite BAG groups, with progressively worse survival among participants with higher BAGs (high versus low: HR, 1.77; P = 1.0 × 10^−45^; high versus average: HR, 1.45; P = 5.2 × 10^−32^; **Fig. 4b**).

**Fig. 4:**
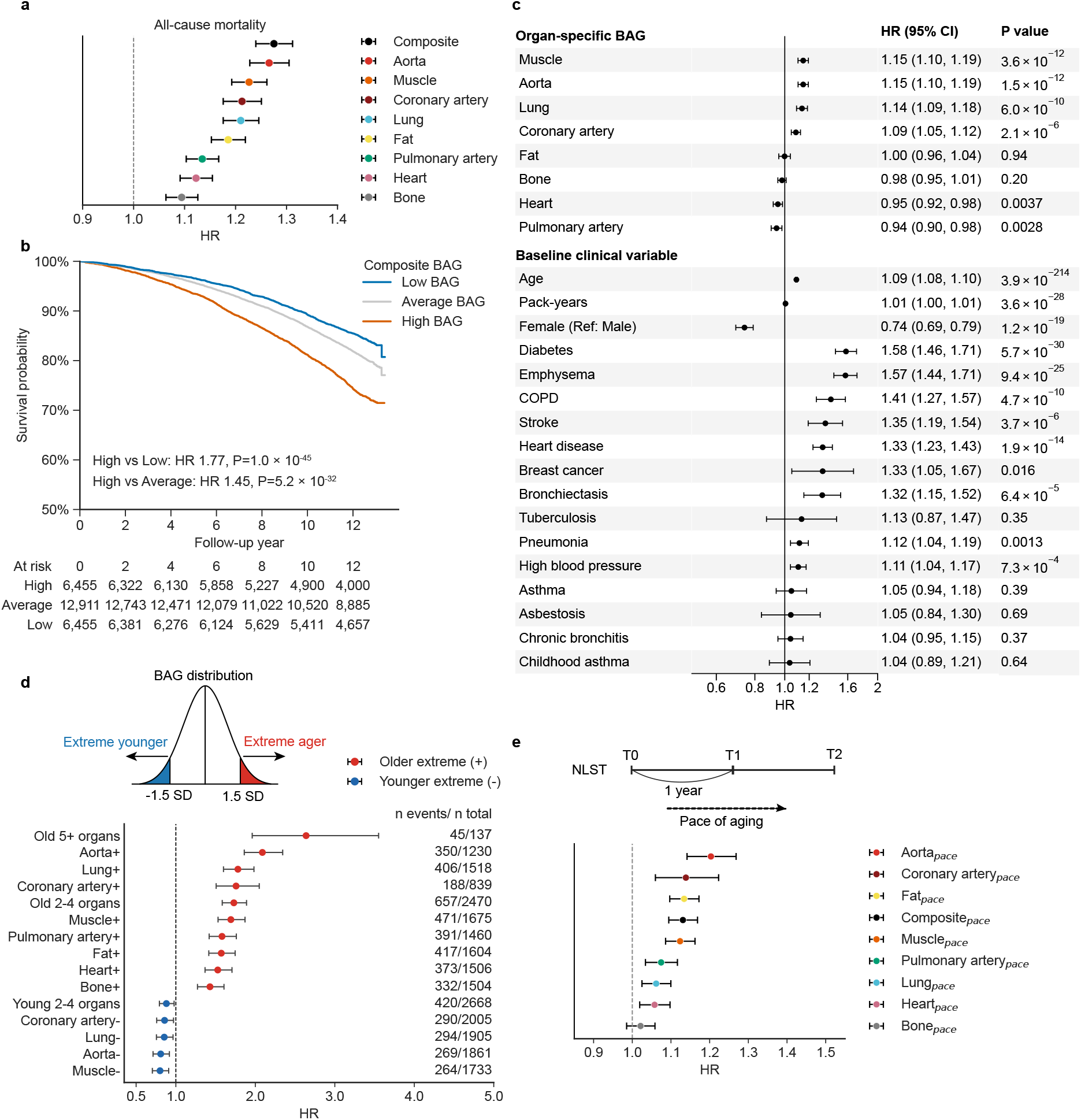
Associations between CT-derived organ aging and all-cause mortality. **a**, Associations between CT-derived BAGs and all-cause mortality. Forest plots show hazard ratios for death per 1-s.d. increase in each organ-specific and composite BAG. Cox proportional hazards regression models were adjusted for chronological age, sex, and pack-years. **b**, All-cause mortality stratified by composite BAG in NLST. Kaplan–Meier curves are shown for participants with low, average, and high composite BAG. HRs for high versus low and high versus average BAG categories were estimated using Cox proportional hazards models adjusted for chronological age, sex, and pack-years. **c**, Organ-specific aging measures independently associated with all-cause mortality. Multivariable Cox models included all eight organ-specific BAGs simultaneously and were adjusted for baseline comorbidities, chronological age, sex, and pack-years. **d**, Mortality risk according to extreme CT-derived organ aging. Participants were classified as extreme agers or extremely youthful agers if their BAG was more than 1.5 s.d. above or below the cohort mean, respectively; only significant associations are shown. **e**, Associations between organ-specific pace of aging and all-cause mortality. Pace was estimated from serial CT scans, and Cox models were adjusted for chronological age, sex, pack-years, and the corresponding baseline BAG. For forest plots in **a**, **c, d**, and **e**, point estimates indicate hazard ratios, and horizontal lines indicate 95% confidence intervals. BAG, biological age gap. COPD, chronic obstructive pulmonary disease.

To determine organ-specific aging measures independently associated with all-cause mortality, we fitted multivariable Cox models that included all eight organ-specific BAGs and baseline comorbidities in addition to chronological age, sex, and pack-years. Muscle, aorta, lung, and coronary artery BAGs remained independently associated with mortality (HR per 1-s.d. higher BAG: muscle BAG, HR, 1.15; P = 3.6 × 10^−12^; aorta BAG, HR, 1.15; P = 1.5 × 10^−12^; lung BAG, HR, 1.14; P = 6.0 × 10^−10^; coronary artery BAG, HR, 1.09; P = 2.1 × 10^−6^) (**Fig. 4c**).

Cause-specific mortality analyses were then performed for four prespecified categories: respiratory, cardiovascular, cancer, and other mortality (**Extended Data Fig. 4**). For respiratory and cardiovascular mortality, BAGs in the corresponding organ systems showed the largest hazard ratios. Lung BAG showed the largest association with respiratory mortality (HR per 1-s.d. higher BAG, 1.47; q = 2.1 × 10^−21^), whereas coronary artery BAG showed the largest observed association with cardiovascular mortality (HR, 1.45; q = 1.1 × 10^−25^) followed by aorta BAG (HR, 1.44; q = 6.6 × 10^−27^). For cancer mortality, the composite BAG showed the largest observed association (HR, 1.15; q = 7.2 × 10^−10^), whereas aorta BAG showed the largest observed association among organ-specific BAGs in these models (HR, 1.15; q = 6.1 × 10^−9^).

Across the four cause-specific mortality categories, aorta BAG was consistently among the three BAGs with the highest observed HRs, highlighting vascular aging as a consistent predictor of mortality across diverse causes of death^37,38^.

### Extreme multi-organ aging burden identifies individuals at highest mortality risk

Building on prior aging-clock studies showing that extreme biological aging identifies accelerated or youthful aging subgroups^9,32^, we examined whether extreme organ-specific BAGs by MOSAIC-Age were associated with mortality. For each organ, participants with BAGs more than 1.5 s.d. above or below the cohort mean were classified as having extremely aged or extremely youthful organs, respectively. Participants with extremely aged organs had a higher hazard of all-cause mortality across multiple organ systems; among the significant associations, the largest HRs were observed for aorta (HR, 2.09; q = 1.5 × 10^−34^), lung (HR, 1.78; q = 4.6 × 10^−25^), and coronary artery (HR, 1.76; q = 1.6 × 10^−12^). Conversely, extremely youthful BAG profiles in muscle, aorta, lung, and coronary artery were associated with reduced mortality risk (muscle: HR, 0.80; q = 1.2 × 10^−3^; aorta: HR, 0.81; q = 1.8 × 10^−3^; lung: HR, 0.85; q = 0.017; coronary artery: HR, 0.86; q = 0.021) (**Fig. 4d**).

Mortality risk increased progressively with greater multi-organ aging burden (**Fig. 4d**). Compared with participants with neither extremely aged nor extremely young organs, higher hazards of all-cause mortality were observed for those with 2–4 extremely aged organs (HR, 1.73; q = 5.2 × 10^−33^) and ≥ 5 extremely aged organs (HR, 2.64; q = 3.4 × 10^−10^). Similar patterns were observed across cause-specific analyses (**Extended Data Fig. 5**).

### Accelerated pace of organ aging predicts mortality

Finally, we investigated whether longitudinal changes in organ-specific BAGs were associated with mortality risk. Using serial CT scans from the NLST trial, we estimated organ-specific pace of aging using linear mixed-effects models. We initiated follow-up from the final CT scan used for pace estimation to mitigate immortal-time bias. After adjustment for chronological age, sex, smoking pack-years, and the corresponding baseline BAG, faster pace of aging was independently associated with higher mortality for all organs except bone (**Fig. 4e**). The largest association was observed for aorta pace (HR per 1-s.d. faster pace, 1.20; q = 1.7 × 10^−11^), indicating that accelerated vascular aging is a particularly strong predictor of mortality risk.

Cause-specific mortality analyses showed similarly strong associations for aortic aging pace across multiple categories (**Extended Data Fig. 6**). Aorta pace had the largest estimates for respiratory mortality (HR per 1-s.d. faster pace, 1.29; q = 8.7 × 10^−4^), cardiovascular mortality (HR, 1.29; q = 9.3 × 10^−5^), and cancer mortality (HR, 1.17; q = 2.6 × 10^−4^). Together, these findings demonstrate that both baseline organ-specific aging and its longitudinal progression pace provide prognostic information for mortality risk.

## Discussion

In this multicohort analysis, we developed MOSAIC-Age, a framework for estimating organ-specific biological age from routine chest CT, using 9,971 examinations from 6,912 individuals without documented abnormalities. We then applied the framework to two independent prospective cohorts including 35,293 participants with longitudinal follow-up. To our knowledge, this represents the largest multicohort study of CT-derived organ aging and its associations with longitudinal health outcomes in tobacco-exposed populations. CT-derived BAGs were associated with lifestyle, socioeconomic, and environmental factors; prevalent comorbidities; future chronic disease risks; and all-cause and cause-specific mortality. Mortality risk also increased progressively with both the number of extremely aged organs and faster aging pace. Together, these findings demonstrate that routine chest CT contains anatomically localized, age-related information associated with clinically important outcomes after accounting for chronological age.

Our findings support a view of biological aging as a spatially heterogeneous and temporally dynamic process. The modest correlations among organ-specific BAGs indicate that aging is not expressed uniformly across the body, while the graded mortality risk associated with the number of extremely aged organs suggests that the accumulation of organ-level deficits may capture a transition from localized vulnerability to broader multisystem decline. Serial imaging further shows that these profiles are not static: organs within the same individual may age at different rates, and these trajectories carry prognostic information beyond their baseline biological age. MOSAIC-Age therefore extends biological age assessment beyond a single whole-person estimate to an anatomically resolved profile of which organs appear most vulnerable and how their aging trajectories evolve.

These findings also broaden the information that can be extracted from routinely acquired chest CT. Although chest CT is generally obtained to address a specific clinical question, such as lung cancer screening or evaluation of cardiopulmonary disease, the same scan offers a window for opportunistic assessment of current physiological conditions and future risks. For example, incident lung cancer was associated not only with lung BAG but also with aorta, coronary artery, fat, and muscle BAGs, indicating that CT correlates of lung cancer risk extend beyond the lung itself. Consistent with this concept, a recent study linked CT-derived thymic health to future lung cancer and cardiovascular outcomes, even though the thymus is not conventionally assessed for either risk^39^. Together, these observations illustrate how an already acquired scan may be repurposed to assess biological aging across multiple organ systems without additional image acquisition. By integrating organ-specific aging signals, CT-derived BAGs could complement conventional risk factors and identify individuals whose vulnerability is not captured by chronological age or single-organ measures. Potential applications include identifying individuals who may benefit from lung cancer interception or cardiometabolic assessment during lung cancer screening and recognizing multisystem vulnerability not captured by conventional risk factors^40^. Because the present analyses were observational, prospective studies must determine whether these measures provide incremental clinical utility, inform actionable interventions and improve patient outcomes.

Longitudinal imaging adds a temporal dimension to this opportunistic assessment. A single examination identifies organs that appear older or younger than expected, whereas serial examinations reveal how these deviations evolve. BAG pace measurements provide a means to distinguish an organ that appears old but remains stable from one undergoing rapid structural aging – two states that may carry different biological and clinical implications. In our study, a faster organ-specific aging pace was associated with higher mortality after adjustment for the corresponding baseline BAG, indicating that temporal change provides prognostic information beyond a single-time-point estimate. This observation is consistent with previous longitudinal studies showing that changes in biological age capture aging dynamics not reflected by single-time-point estimates^41^. Complementing our findings, recent population-based evidence has linked accelerated biological aging measured from blood to an increased risk of early onset of cancers^42^. In patients already undergoing serial chest CT for cancer screening or disease-specific follow-up, organ-specific BAG trajectories warrant evaluation as biomarkers of biological change during or following preventive or therapeutic interventions.

CT-derived aging clocks are complementary to blood-based aging clocks. Proteomic aging clocks infer organ-specific aging from circulating molecular signatures^9,10,32,43^, where CT directly localizes age-related structural manifestations within individual organs. These manifestations may reflect the cumulative structural effects of tissue remodeling, ectopic fat deposition, vascular change, muscle loss, chronic inflammation and environmental exposures. MOSAIC-Age does not distinguish these mechanisms directly, but its anatomical resolution provides a framework for linking molecular aging processes to their organ-level structural consequences. MRI-based clocks characterize structural aging across multiple organs^36^, but provide limited assessment of the lungs and are not routinely used in tobacco-exposed populations. MOSAIC-Age adds an anatomically resolved thoracic perspective by quantifying organ morphology and tissue composition on routinely acquired chest CT. Given that tens of millions of chest CT examinations are performed worldwide annually, this approach could enable opportunistic assessment at substantial scale without additional cost. Integrating these modalities may better characterize biological aging across molecular and structural domains.

Because CT entails exposure to ionizing radiation, a completely healthy reference population cannot readily be assembled through dedicated imaging. We therefore used two large, independent CT datasets drawn from general populations on two continents and systematically reviewed available radiology reports to exclude examinations with documented abnormalities. The datasets encompassed heterogeneous acquisition protocols, scanner platforms and reconstruction parameters, allowing model performance to be evaluated across technical variations encountered in clinical practice. MOSAIC-Age was evaluated in held-out samples from both cohorts and further validated in an independent cohort from our cancer center. Although undocumented subclinical abnormalities could not be completely excluded, the anatomical concordance between organ-specific BAGs and related clinical outcomes supports their biological relevance. Accordingly, BAGs should be interpreted as relative measures of organ appearance on CT rather than direct measures of disease-free aging.

Among the organ-specific measures, aorta BAG showed the strongest observed associations with both incident lung cancer and cancer mortality. Rather than indicating a causal effect of local aortic aging on carcinogenesis, this association may reflect shared risk factors and biological processes underlying vascular disease and cancer, including smoking, chronic inflammation, oxidative stress and metabolic dysregulation^44^. Supporting an inflammatory link, the CANTOS trial showed that inhibition of interleukin-1β reduced cardiovascular events among patients with atherosclerosis and persistent inflammation and, in exploratory analyses, was associated with dose-dependent reductions in lung cancer incidence and mortality^45^. Smoking represents another shared contributor: greater smoking exposure was associated with all organ-specific BAGs in our evaluation cohorts, and NLST enrolled only current and former smokers. As a CT-derived measure, aorta BAG likely represents a structural phenotype shaped by vascular aging, cumulative exposures and systemic biological processes, rather than a readout of any single pathway. Its association with cancer outcomes may therefore identify a systemic phenotype relevant to both vascular disease and cancer. The biological mechanisms and potential causal relationships underlying this association warrant investigation in future mechanistic studies.

These findings define several priorities for future investigation. Evaluation in broader populations, particularly never-smokers and individuals undergoing chest CT for indications beyond lung disease, will be important for establishing generalizability. Prospective studies should determine whether organ-specific BAGs add clinically meaningful information beyond established risk factors and whether their longitudinal trajectories are modifiable through preventive or therapeutic interventions. Integration with molecular, clinical and environmental measurements may further clarify the biological processes underlying CT-derived organ aging and distinguish age-related structural variation from subclinical disease and cumulative exposure. Together, these studies will determine whether CT-derived aging profiles can support personalized monitoring and risk-adapted prevention in routine clinical care.

In conclusion, chest CT-derived organ aging clocks capture clinically meaningful age-related structural variation beyond chronological age. Organ-specific BAGs were associated with lifestyle factors, chronic disease, and mortality. By providing a structural imaging readout of multi-organ aging, routine chest CT may offer new opportunities for opportunistic health assessment and personalized risk stratification.

## Author contributions

J.S. and J.W. conceived the clinical problem and contributed to the overall study design. J.S., M.S., A.Z., and J.W. developed and trained the models and conducted the statistical analyses. J.S., A.M., X.X., and E.Z. performed data quality checks. J.S. prepared the draft. N.I.V., T.C., X.L., M.A., E.E.G., A.S., E.J.O., A.A.S., T.L., M.M., A.A.C., D.E.G., F.U.K., M.C.B.G., B.W.C., G.S.S., L.A.B., C.C., D.J., D.R., Z.L., J.Y.C., A.A.V., D.L.G., C.C.W., J.V.H., J.Z., and J.W. critically reviewed and edited the manuscript. All authors reviewed and approved the final version.

## Acknowledgements

This research was partially supported by NIH grants R01CA262425 and R01CA276178, as well as CPRIT RP240117. This work was supported by Victory Houston, Permanent Health Funds, and QIAC Partnership in Research Grant. Furthermore, this work was supported by generous philanthropic contributions from Andrea Mugnaini and Edward L. C. Smith. Finally, this work was supported by Rexanna’s Foundation for Fighting Lung Cancer. The funding sources had no role in the study design; data collection, analysis, or interpretation; or manuscript preparation. We thank the participants, investigators, data contributors, and teams responsible for creating and maintaining CT-RATE, MIDRC, NLST, and COPDGene, whose collective efforts made this study possible. Full acknowledgements for each dataset are provided in the **Supplementary Note 1**.

## Extended Data

**Extended Data Fig. 1.**
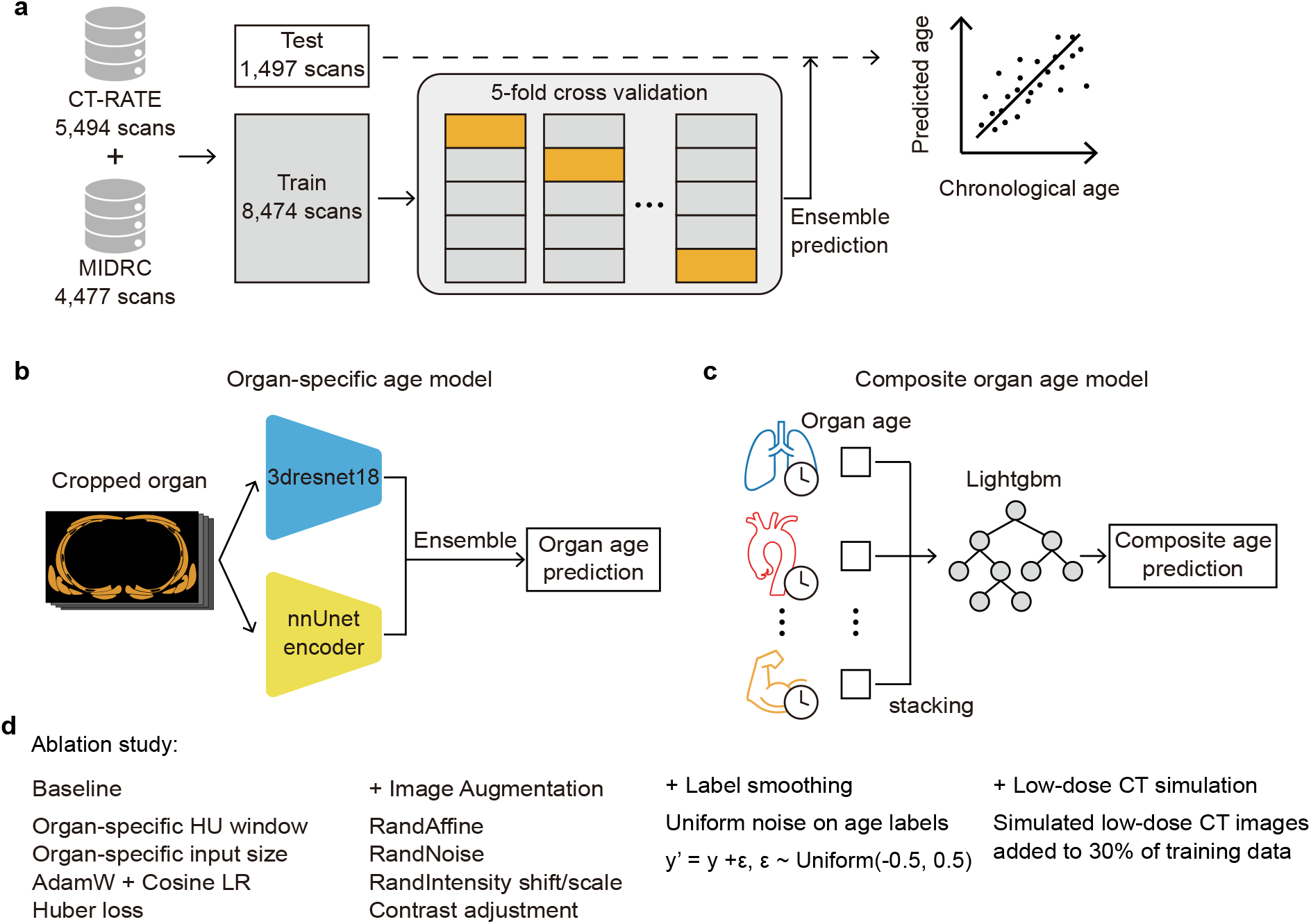
Development and evaluation workflow for CT-based organ aging clocks. **a**, Model development and evaluation strategy. The CT-RATE and MIDRC datasets were used to develop the organ aging clocks. The combined dataset was split into a held-out test set of 1,497 scans (15%) and a training set of 8,474 scans (85%) without patient overlap. Five-fold cross-validation was performed within the training set, and ensemble predictions from the five trained models were evaluated in the held-out test set by comparing predicted age with chronological age. **b**, Organ-specific age model architecture. Each organ-specific age model used two encoder networks, a 3D ResNet-18 and an nnU-Net encoder, whose outputs were combined to generate the final organ age prediction. **c**, Composite organ age model. Organ-specific age predictions were integrated using LightGBM-based stacking to generate the composite age prediction. **d**, Components evaluated in the ablation study. The ablation study assessed the contribution of training strategies, data augmentation, label smoothing, and low-dose CT simulation to model performance.

**Extended Data Fig. 2.**
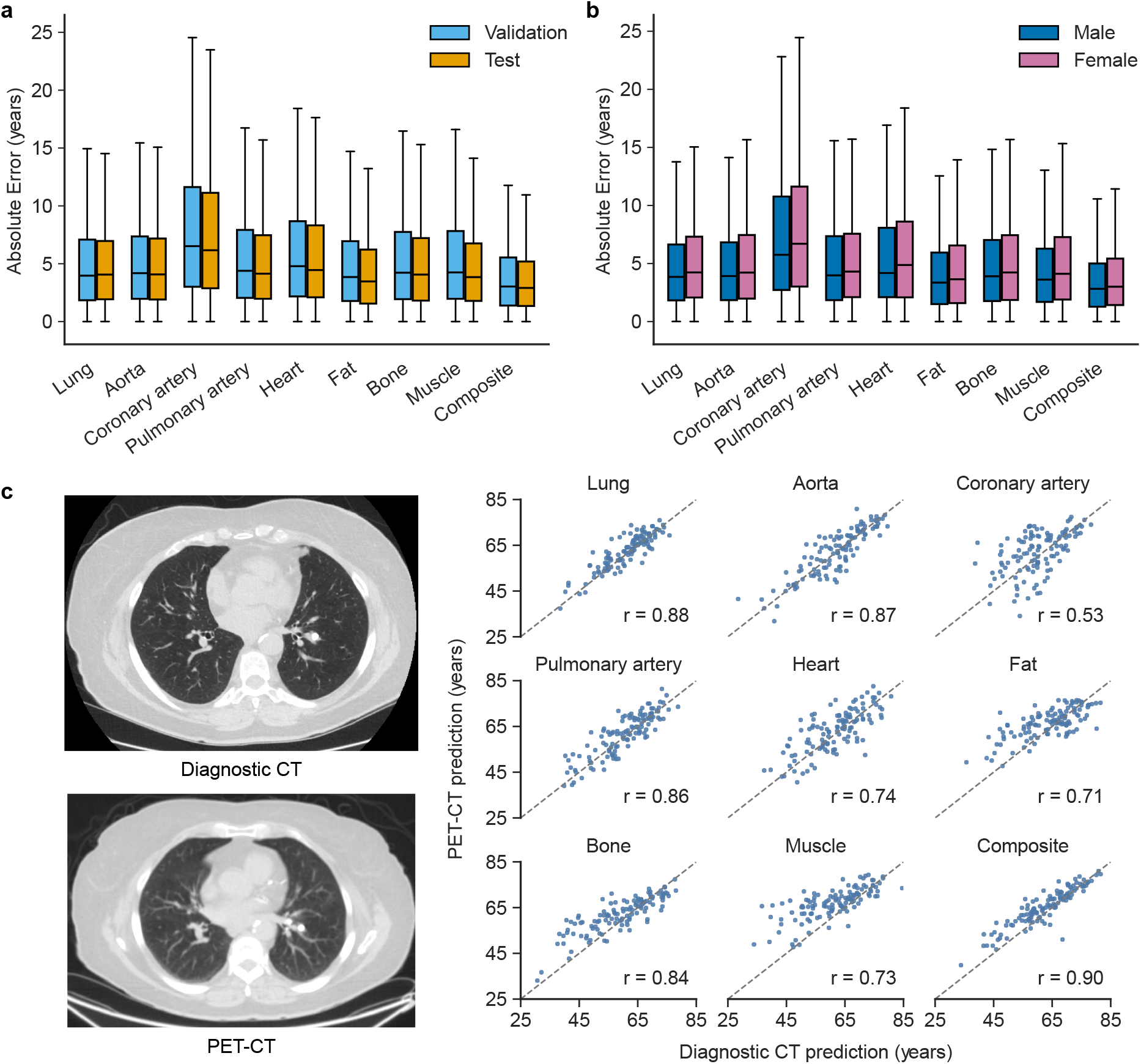
Validation analyses of CT-based organ aging clocks. **a**, Absolute error distributions for organ-specific and composite aging clocks in the validation and held-out test datasets. **b**, Absolute error distributions by sex for organ-specific and composite aging clocks. **c**, Agreement between age predictions derived from diagnostic CT and the CT component of PET/CT among participants with paired examinations acquired within a similar time window. Representative paired CT images are shown on the left, with scatter plots on the right comparing age predictions for each organ-specific and composite aging clock; Pearson’s correlation coefficients are reported in each plot. For box plots, center lines show medians, boxes show IQR, and whiskers extend to values within 1.5 × IQR.

**Extended Data Fig. 3.**
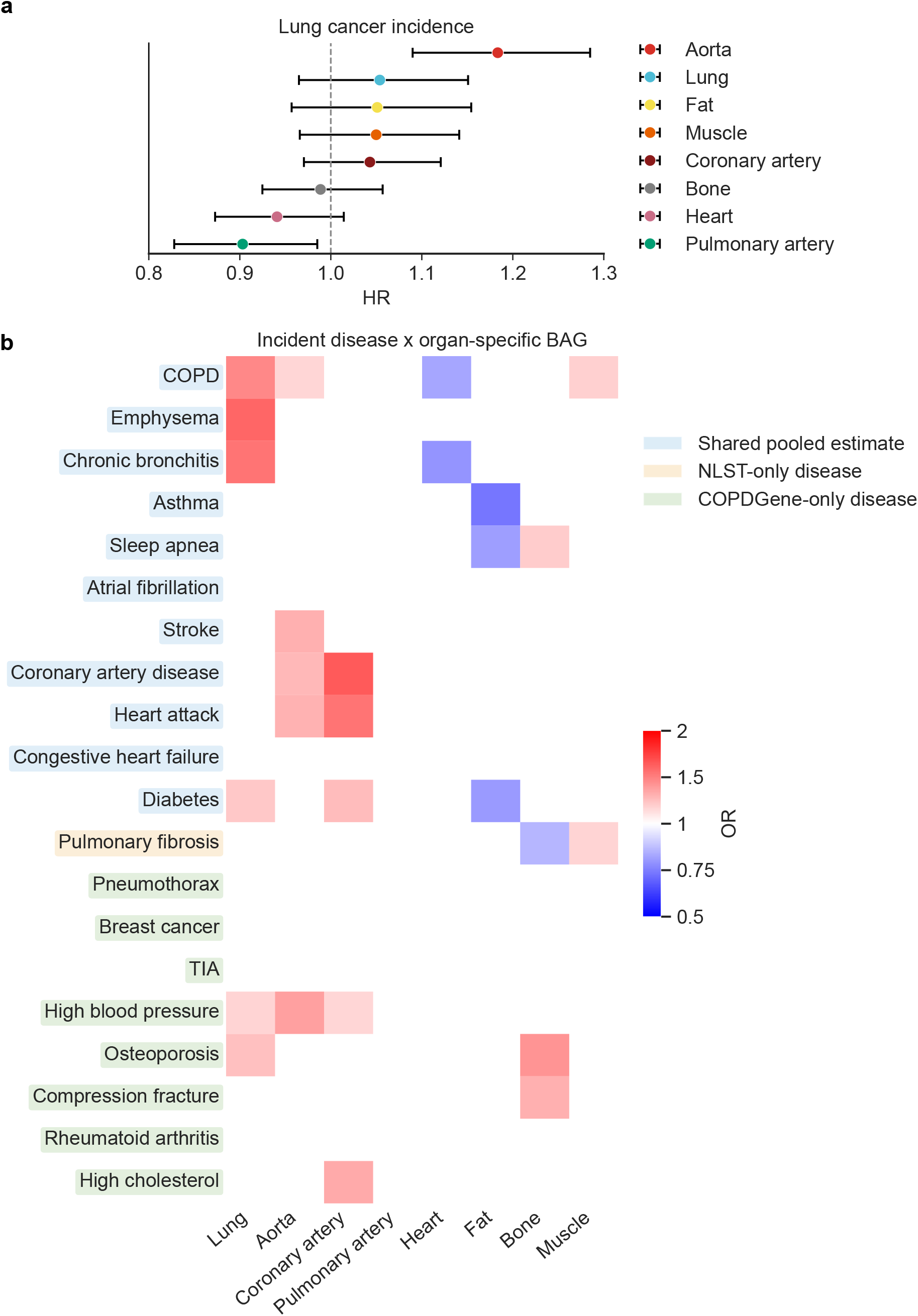
Mutually adjusted associations between CT-derived BAGs and incident disease. **a**, Mutually adjusted associations between organ-specific BAGs and incident lung cancer. **b**, Mutually adjusted associations between BAGs and 20 incident chronic disease outcomes. **a**,**b**, BAGs were mutually adjusted by entering all relevant BAGs simultaneously into the same model. Cox proportional hazards regression was used for incident lung cancer in **a**, and logistic regression was used for incident chronic disease in **b**. Models were adjusted for chronological age, sex, and pack-years. In **a**, point estimates indicate hazard ratios, and horizontal lines indicate 95% confidence intervals. In **b**, heatmap colors indicate odds ratios; only associations with q < 0.05 are color coded. BAG, biological age gap; COPD, chronic obstructive pulmonary disease; TIA, transient ischemic attack.

**Extended Data Fig. 4.**
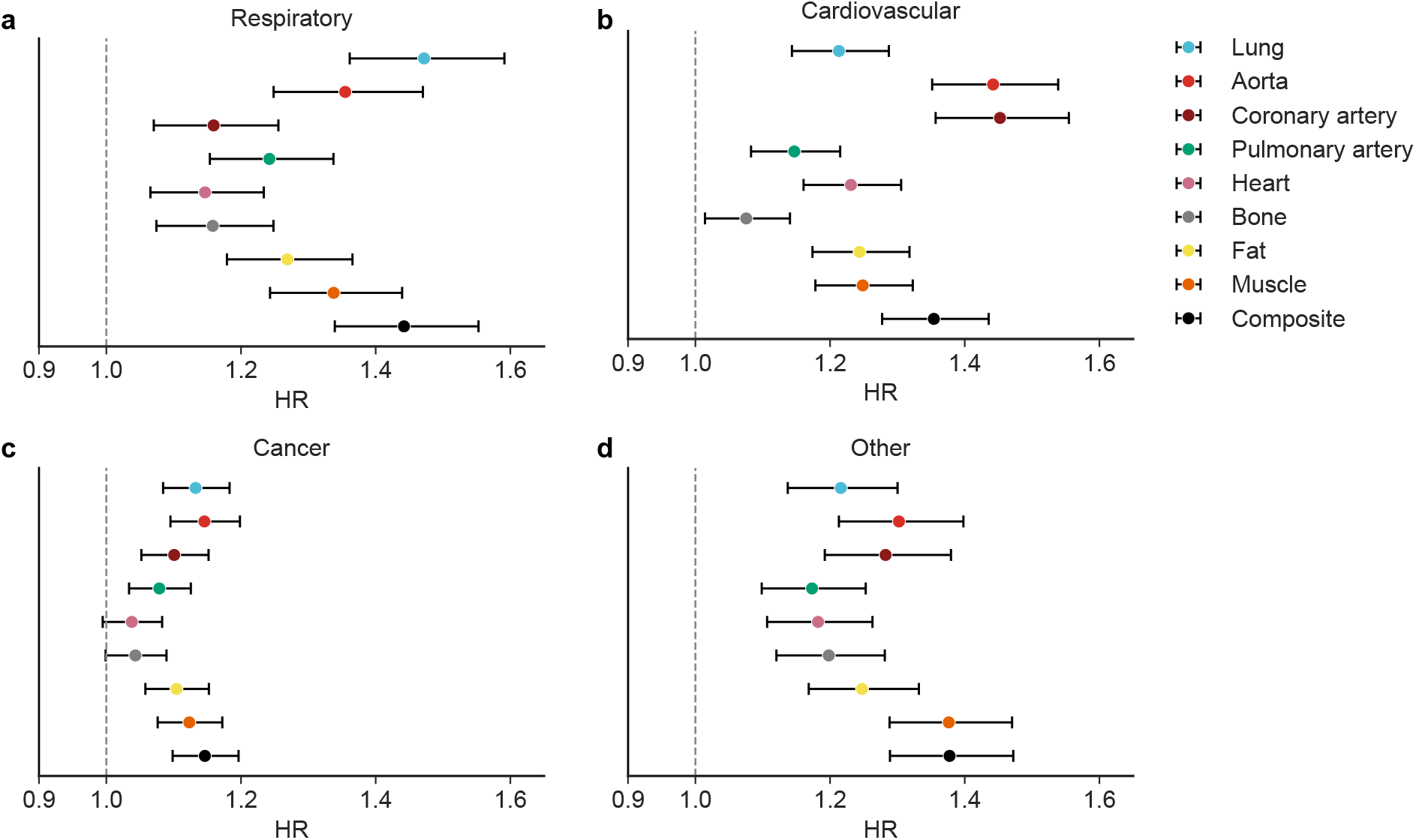
Associations between CT-derived BAGs and cause-specific mortality. Forest plots show hazard ratios for cause-specific mortality per 1-s.d. increase in each organ-specific and composite BAG, estimated using Cox proportional hazards regression adjusted for chronological age, sex, and pack-years. Point estimates indicate hazard ratios, and horizontal lines indicate 95% confidence intervals. **a**, Respiratory mortality. **b**, Cardiovascular mortality. **c**, Cancer-related mortality. **d**, Other-cause mortality. BAG, biological age gap; HR, hazard ratio.

**Extended Data Fig. 5.**
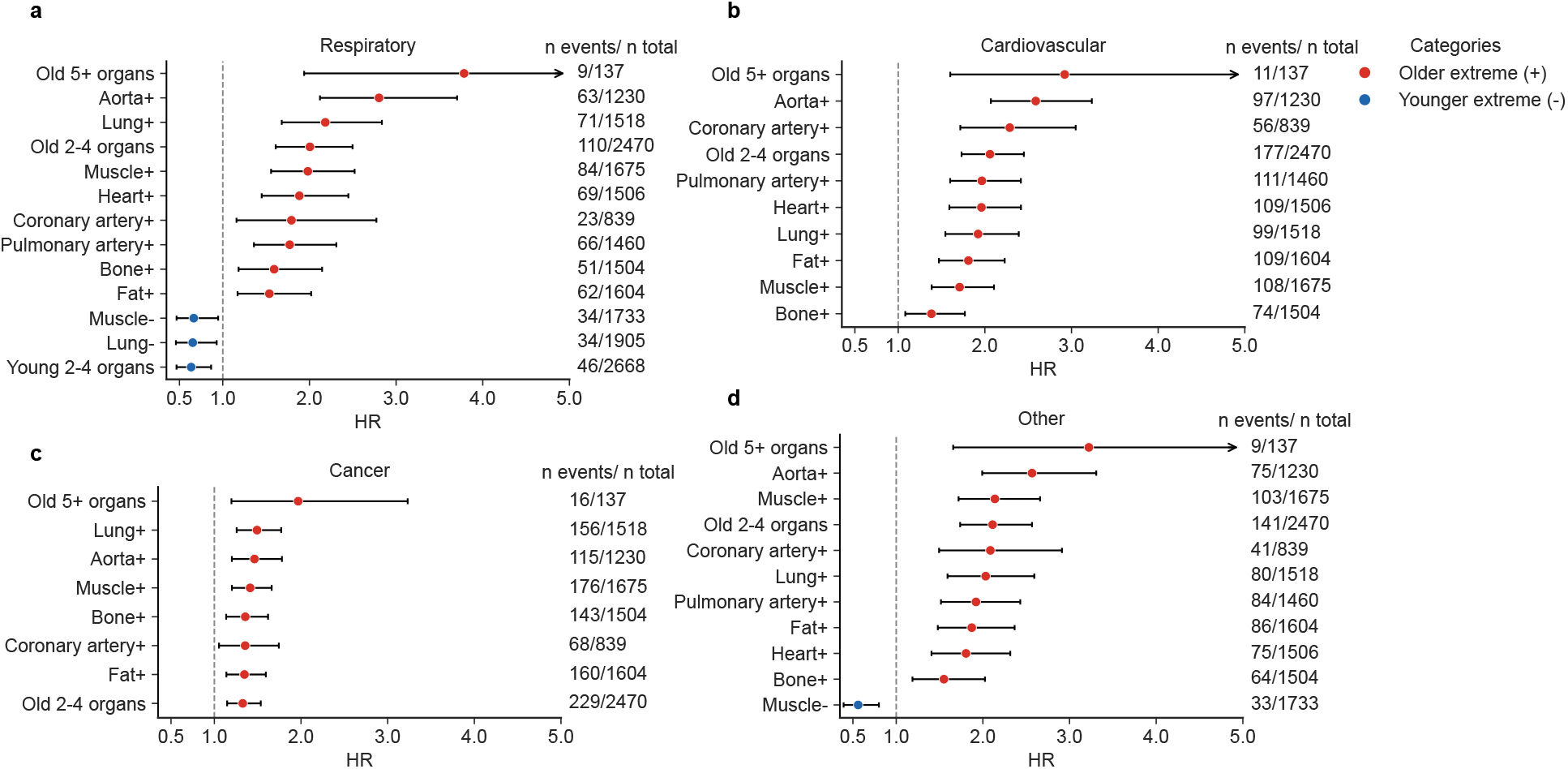
Associations of CT-derived extreme organ aging with cause-specific mortality. Forest plots show associations of extreme organ aging with cause-specific mortality. For extreme organ aging, participants were classified as extreme agers or extremely youthful agers if their BAG was more than 1.5 s.d. above or below the cohort mean, respectively; only significant associations are shown. Point estimates indicate hazard ratios, and horizontal lines indicate 95% confidence intervals. Arrows indicate 95% confidence intervals extending beyond the plotted x-axis limits. **a**, Respiratory mortality. **b**, Cardiovascular mortality. **c**, Cancer-related mortality. **d**, Other-cause mortality. BAG, biological age gap; CI, confidence interval; HR, hazard ratio.

**Extended Data Fig. 6.**
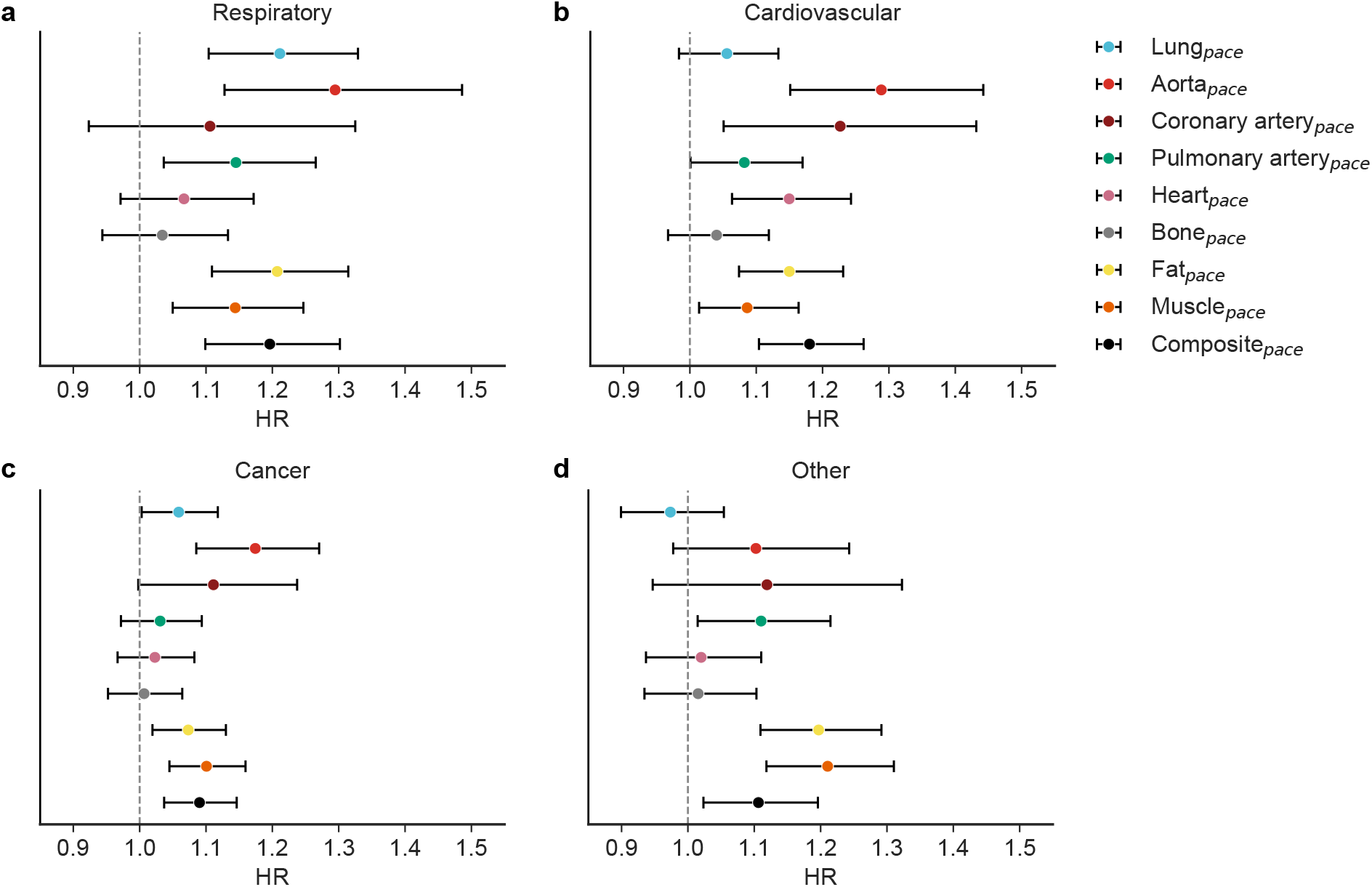
Associations of CT-derived aging pace with cause-specific mortality. Forest plots show associations of organ-specific BAG pace with cause-specific mortality. Hazard ratios are shown per 1-s.d. increase and were estimated using Cox models adjusted for chronological age, sex, pack-years, and baseline BAG. Point estimates indicate hazard ratios, and horizontal lines indicate 95% confidence intervals. **a**, Respiratory mortality. **b**, Cardiovascular mortality. **c**, Cancer-related mortality. **d**, Other-cause mortality. BAG, biological age gap; CI, confidence interval; HR, hazard ratio.

**Extended Data Table. 1.** Ablation study of CT-derived organ aging clock development.

| Model | IT | LS | LT | Lung | Aorta | Coronary artery | Pulmonary artery | Heart | Bone | Fat | Muscle | Composite | Average |
| --- | --- | --- | --- | --- | --- | --- | --- | --- | --- | --- | --- | --- | --- |
|  |  |  |  | 5.36 ± 0.25 | 5.27 ± 0.14 | 8.76 ± 0.18 | 5.55 ± 0.13 | 6.36 ± 0.16 | 5.34 ± 0.06 | 4.92 ± 0.18 | 5.05 ± 0.21 | 4.03 ± 0.12 | 5.63 |
|  | ✓ |  |  | 5.16 ± 0.15 | 5.21 ± 0.13 | 8.58 ± 0.22 | 5.53 ± 0.11 | 6.18 ± 0.09 | 5.20 ± 0.14 | 4.95 ± 0.05 | 4.84 ± 0.21 | **3.94 ± 0.12** | 5.51 |
| **nnU-E** |  | ✓ |  | 5.39 ± 0.17 | 5.29 ± 0.07 | 8.82 ± 0.12 | 5.59 ± 0.15 | 6.28 ± 0.10 | 5.31 ± 0.12 | 4.84 ± 0.10 | 4.95 ± 0.18 | 4.01 ± 0.11 | 5.61 |
|  |  |  | ✓ | 5.39 ± 0.22 | 5.28 ± 0.15 | 8.81 ± 0.24 | 5.61 ± 0.12 | 6.13 ± 0.03 | 5.23 ± 0.18 | 4.78 ± 0.11 | 4.90 ± 0.14 | 3.96 ± 0.11 | 5.57 |
|  | ✓ | ✓ | ✓ | 5.26 ± 0.21 | 5.19 ± 0.15 | 8.47 ± 0.15 | 5.52 ± 0.09 | 6.11 ± 0.13 | **5.11 ± 0.09** | 5.13 ± 0.46 | 5.20 ± 0.53 | 4.05 ± 0.27 | 5.56 |
|  |  |  |  | 5.31 ± 0.11 | 5.47 ± 0.13 | 12.26 ± 6.75 | 5.47 ± 0.06 | 6.17 ± 0.16 | 5.73 ± 0.08 | 4.61 ± 0.12 | 5.23 ± 0.24 | 3.99 ± 0.13 | 6.03 |
|  | ✓ |  |  | 5.17 ± 0.12 | 5.43 ± 0.12 | 8.21 ± 0.10 | 5.56 ± 0.11 | 6.20 ± 0.16 | 5.78 ± 0.16 | 4.59 ± 0.09 | 5.17 ± 0.11 | 4.08 ± 0.12 | 5.58 |
| **R18** |  | ✓ |  | 5.32 ± 0.16 | 5.45 ± 0.15 | 8.91 ± 0.94 | 5.52 ± 0.08 | 6.28 ± 0.16 | 5.62 ± 0.04 | 4.69 ± 0.07 | 5.24 ± 0.05 | 4.05 ± 0.08 | 5.68 |
|  |  |  | ✓ | 5.39 ± 0.13 | 5.48 ± 0.19 | 8.47 ± 0.17 | 5.55 ± 0.11 | 6.30 ± 0.15 | 5.67 ± 0.06 | 4.59 ± 0.08 | 5.22 ± 0.14 | 4.01 ± 0.08 | 5.63 |
|  | ✓ | ✓ | ✓ | 5.21 ± 0.13 | 5.40 ± 0.11 | 8.26 ± 0.13 | 5.52 ± 0.12 | 6.19 ± 0.17 | 5.74 ± 0.06 | 4.73 ± 0.27 | 5.34 ± 0.35 | 4.14 ± 0.23 | 5.61 |
|  |  |  |  | 5.26 ± 0.20 | 5.23 ± 0.16 | 9.87 ± 2.59 | **5.37 ± 0.08** | 6.04 ± 0.15 | 5.36 ± 0.03 | 4.55 ± 0.07 | 4.90 ± 0.17 | 4.39 ± 0.22 | 5.66 |
|  | ✓ |  |  | **5.10 ± 0.14** | 5.19 ± 0.15 | 8.21 ± 0.11 | 5.41 ± 0.10 | 6.02 ± 0.14 | 5.30 ± 0.13 | 4.56 ± 0.07 | **4.80 ± 0.16** | 4.44 ± 0.29 | 5.45 |
| **ENS** |  | ✓ |  | 5.27 ± 0.17 | 5.25 ± 0.11 | 8.66 ± 0.36 | 5.43 ± 0.04 | 6.07 ± 0.16 | 5.29 ± 0.10 | 4.54 ± 0.07 | 4.85 ± 0.09 | 4.30 ± 0.14 | 5.52 |
|  |  |  | ✓ | 5.32 ± 0.18 | 5.25 ± 0.18 | 8.41 ± 0.09 | 5.43 ± 0.09 | 6.03 ± 0.09 | 5.25 ± 0.08 | **4.51 ± 0.07** | 4.84 ± 0.11 | 4.22 ± 0.24 | 5.47 |
|  | ✓ | ✓ | ✓ | 5.18 ± 0.18 | **5.17 ± 0.12** | **8.19 ± 0.06** | 5.37 ± 0.10 | **5.98 ± 0.14** | 5.22 ± 0.06 | 4.56 ± 0.20 | 4.95 ± 0.32 | 4.36 ± 0.26 | **5.44** |

Data are mean absolute errors in years, presented as mean ± SD. Average is the mean of the eight organ-specific and composite values and is shown without SD. Check marks indicate the training strategies applied in each configuration. Boldface indicates the lowest value in each column. ENS, ensemble of R18 and nnU-E; IT, image transformation; LS, label smoothing; LT, LDCT transformation; nnU-E, nnU-Net encoder; R18, 3D ResNet-18.

**Extended Data Table. 2.** Baseline characteristics of the NLST and COPDGene cohort.

| Cohort | NLST (N = 25,824) | COPDGene (N = 9,469) |
| --- | --- | --- |
| Analysis cohort, *n* (%) |  |  |
| T1 available among T0 patients | 23,562 (91.2%) | NA |
| T2 available among T0 patients | 22,731 (88.0%) | NA |
| Age, mean (SD) | 61.4 (5.0) | 59.3 (9.0) |
| Female sex, *n* (%) | 10,549 (40.9%) | 4,423 (46.7%) |
| BMI, mean (SD) | 27.9 (5.0) | 28.8 (6.2) |
| Race, *n* (%) |  |  |
| White | 23,588 (91.3%) | 6,456 (68.2%) |
| Black or African-American | 1,104 (4.3%) | 3,013 (31.8%) |
| Asian | 542 (2.1%) | 0 (0.0%) |
| Other/unknown race | 590 (2.3%) | 0 (0.0%) |
| Pack-years, mean (SD) | 56.0 (24.0) | 44.0 (25.2) |

## Data availability

CT-RATE imaging data are available through the Hugging Face repository under the repository’s access conditions for academic, research, and educational use (https://huggingface.co/datasets/ibrahimhamamci/CT-RATE). MIDRC imaging data are available through the MIDRC Data Commons to registered users under the applicable MIDRC data use agreement (https://data.midrc.org/). The NLST imaging data are available through The Cancer Imaging Archive (https://www.cancerimagingarchive.net/collection/nlst/), and NLST clinical data are available through the NCI Cancer Data Access System (https://cdas.cancer.gov/nlst/). The COPDGene phenotype and imaging data used in this study are available through controlled access via dbGaP under study accession numbers phs000179.v7.p2 and phs004023.v1.p2, respectively. The paired CT/PET-CT data are not publicly available owing to patient privacy considerations but are available for research purposes from the corresponding author upon reasonable request. The pretrained organ-clock model weights are also available from the corresponding author upon reasonable request.

## Code availability

Our code for model inference will be publicly available at GitHub (https://github.com/WuLabMDA/MOSAIC-Age).

