## Supplementary Information for "Multi-organ aging quantified from routine chest CT predicts chronic disease risk and mortality"

### Contents of Supplementary Information

Supplementary Methods
    Supplementary Method 1– page 2,3
    Supplementary Method 2 – page 3,4
    Supplementary Method 3 – page 4
    Supplementary Method 4 – page 4,5
Supplementary Note
    Supplementary Note 1 – page 5
Supplementary Figures
    Supplementary Figure 1 – page 6,7
Supplementary Tables
    Supplementary Table 1 – page 8
    Supplementary Table 2 – page 9
    Supplementary Table 3 – page 10
    Supplementary Table 4 – page 11,12

### Supplementary Methods

**Supplementary Method 1: Organ clock pipeline**

We developed an end-to-end deep-learning-based pipeline to predict chronological age from organ-specific CT images. The pipeline included eight anatomical targets: lung, aorta, coronary arteries, pulmonary artery, heart, bone, fat, and muscle. Each CT image was first processed using a pretrained multi-organ segmentation model. After organ-specific masks were generated, each organ was cropped from the original CT image and used as input to the corresponding organ-clock model. To integrate aging information across organs, we also developed a composite model that integrated the predictions from the organ-specific clocks. The details of the pipeline components and training parameters are described below.

**1.1 Organ segmentation and image processing**

A total of 9,971 chest CT examinations from 6,912 individuals across the MIDRC and CT-RATE datasets were first processed with TotalSegmentator. To extract organ-specific cropped images, we used the following six TotalSegmentator tasks across the target organs: total, heartchambers_highres, coronary_arteries, lung_vessels, vertebrae_body, and tissue_4_types. Using the corresponding segmentation labels, we extracted organ-specific 3D crops from the original CT images. The cropped images were resized and normalized using organ-specific Hounsfield unit (HU) windows to account for differences in organ size and tissue density. For the lung, the five lobes were extracted separately using the same parameters shown below.

| Organ | Totalsegmentator task<br>(task name, label id) | input size<br>(depth, height, width) | HU window<br>(WL, WW) |
|---|---|---|---|
| Lung | (total, 10-14) | (64, 128, 128) | (-500, 1400) |
| Aorta | (heartchambers_highres, 6) | (96, 112, 80) | (75, 550) |
| Coronary artery | (coronary_arteries, 1) | (64, 128, 128) | (75, 550) |
| Pulmonary artery | (lung_vessels, 3) | (128, 192, 192) | (75, 550) |
| Heart | (heartchambers_highres, 1-5) | (64, 128, 128) | (75, 550) |
| Bone | (vertebrae_body, 1,2) | (256, 112, 96) | (500, 1600) |
| Fat | (tissue_4_types, 1,2,4) | (192, 192, 192) | (0, 500) |
| Muscle | (tissue_4_types, 3) | (192, 192, 192) | (75, 550) |

**1.2 Organ clock training**

We developed deep-learning models to automatically predict chronological age from the cropped CT images. After comprehensive ablation experiments across different model architectures and training strategies (see Section 2, Ablation study), we selected the final method based on prediction performance. The final model was an ensemble of two encoders: 3D ResNet-18[1] and an nnU-Net[2] encoder. The 3D ResNet-18 encoder was pretrained on the Kinetics-400 dataset[3], and the nnU-Net encoder was pretrained on the TotalSegmentator dataset[4]. For each encoder, the output feature map was average-pooled and fed into a two-layer fully connected head for age prediction. The CT-RATE and MIDRC datasets were split at the patient level into an 85% training set and a 15% test set, with no patient overlap. Within the training set, we performed five-fold stratified cross-validation. This resulted in five trained models for each organ and each encoder. All organ models were trained with a batch size of 16 for up to 80 epochs. Huber loss was used as the training loss, and mean absolute error (MAE) was used as the evaluation metric. For each fold, the checkpoint with the lowest validation MAE was selected for evaluation. Training was stopped early if the validation MAE did not improve for more than 30 epochs. For the lung, separate models were trained for each of the five lung lobes.

The composite model was trained using LightGBM. For this model, the eight organ-specific age predictions were used as input features to generate the final composite age prediction (**Extended Data Fig. 1b**).

Model training was performed using one to four NVIDIA A100 GPUs, depending on the organ-specific input size. Additional training details are available at: https://github.com/WuLabMDA/MOSAIC-Age

**Supplementary Method 2: Ablation study**

To identify the best model architecture for CT-based age prediction, we compared three deep-learning configurations: 3D ResNet-18 alone, the nnU-Net encoder alone, and an ensemble of the two models. We also tested the following three training strategies and compared their prediction performance.

**Image transform**

In addition to the standard resizing and normalization described in **Supplementary Methods 1.1**, additional image augmentation was applied during training to improve model generalizability. These augmentations included random affine transforms (rotation ±10°, scale ±10%, shear ±10%), random Gaussian noise (mean 0, standard deviation 0.1), random intensity shift (offset ±0.1), random intensity scaling (factor ±0.1), and random contrast adjustment (gamma range 0.7–1.5). Each augmentation was applied independently with a probability of 10%.

**Label smoothing**

The chronological age labels in the training data were recorded as integer values. Therefore, they only approximated the underlying continuous age of each individual. To account for this discretization, we applied label smoothing during training by adding small random noise (±0.5 years) to the chronological age labels.

**LDCT transform**

To improve robustness to variations in radiation dose and image quality across CT scans, we introduced low-dose CT (LDCT)-based augmentation. Using the ASTRA Toolbox[5], standard-dose CT images were forward-projected to generate sinograms. Low-dose conditions were then simulated in the projection domain, and pseudo-LDCT images were reconstructed from the modified sinograms. 30% of the training data were augmented in this manner and used for training.

To reduce the effect of random variation, model performance was evaluated using three train-test splits. For each split, we confirmed that the distributions of age and sex did not differ statistically between the training and test sets. For downstream clinical analyses, predictions were averaged across the models trained using the three train-test splits. The MAE results for all settings are shown in **Extended Data Table 1**.

**Supplementary Method 3: Evaluation of Model Prediction Variability**

**3.1 variability between validation and test set**

To assess model generalizability, we compared the MAE from out-of-fold predictions, defined as predictions on the held-out validation fold during five-fold cross-validation, with the MAE on the separate held-out test set. Predictions from the three train-test splits were pooled to calculate the final MAE.

**3.2 variability between sex**

Previous omics-based multi-organ clock study has reported differences in model outputs by sex[6]. Therefore, we evaluated whether each organ-clock model was affected by sex. In the test set, we calculated and compared the MAE separately for female and male participants.

**3.3 variability between imaging modalities**

To assess the robustness of organ-clock predictions to differences in imaging conditions, we performed a validation analysis using an in-house cohort of 120 patients with early-stage lung cancer, defined as stage I or stage II disease, who had paired diagnostic CT and PET/CT scans acquired during the same period. The diagnostic CT and the CT component of PET/CT differed in radiation dose, field of view, and reconstruction kernel (**Extended Data Fig. 2c**). Because all patients in this cohort had lung cancer, tumor-related findings could affect organ-clock predictions. Therefore, we did not use this cohort to evaluate prediction accuracy against chronological age. Instead, we used the paired scans to assess prediction consistency under different imaging conditions. For each patient, we applied the same organ-clock model to both scans and assessed the correlation between the two predicted ages.

**Supplementary Method 4: Software and reproducibility**

All analyses were performed using Python (v3.12.11). Representative Python packages included torch (v2.4.1), torchvision (v0.19.1), TotalSegmentator (v2.13.0), MONAI (v1.5.2), nnU-Net v2 (v2.6.4), timm (v1.0.26), batchgenerators (v0.25.1), NumPy (v2.4.3), pandas (v2.3.3), scikit-learn (v1.8.0), scikit-image (v0.26.0), SimpleITK (v2.5.3), NiBabel (v5.3.2), ASTRA Toolbox (v2.4.1), PyYAML (v6.0.3), and LightGBM (v4.6.0).

#### Supplementary Note 1: Dataset acknowledgements


##### CT-RATE

We thank the CT-RATE investigators and Istanbul Medipol University Mega Hospital for creating and providing the CT-RATE dataset.

##### MIDRC

The imaging and associated clinical data obtained from the Medical Imaging and Data Resource Center (MIDRC) and used in this study were made possible by the National Institute of Biomedical Imaging and Bioengineering of the National Institutes of Health under contract 75N92020D00021 and through the Advanced Research Projects Agency for Health (ARPA-H). The views and conclusions expressed in this article are those of the authors and should not be interpreted as representing official U.S. Government policy.

##### NLST

We thank the National Cancer Institute (NCI) for access to NCI data collected by the National Lung Screening Trial. The statements contained herein are solely those of the authors and do not represent or imply concurrence or endorsement by the NCI.

##### COPDGene

The COPDGene phenotype and imaging data analyzed in this study were obtained through the NIH database of Genotypes and Phenotypes (dbGaP) under accession numbers phs000179.v7.p2 and phs004023.v1.p2, respectively. We gratefully acknowledge the COPDGene participants, investigators, and data submitters. The COPDGene study (NCT00608764) is supported by grants from the National Heart, Lung, and Blood Institute (U01HL089897 and U01HL089856), by NIH contract 75N92023D00011, and by the COPD Foundation through contributions made to an Industry Advisory Committee that has included AstraZeneca, Bayer Pharmaceuticals, Boehringer-Ingelheim, Genentech, GlaxoSmithKline, Novartis, Pfizer, and Sunovion.

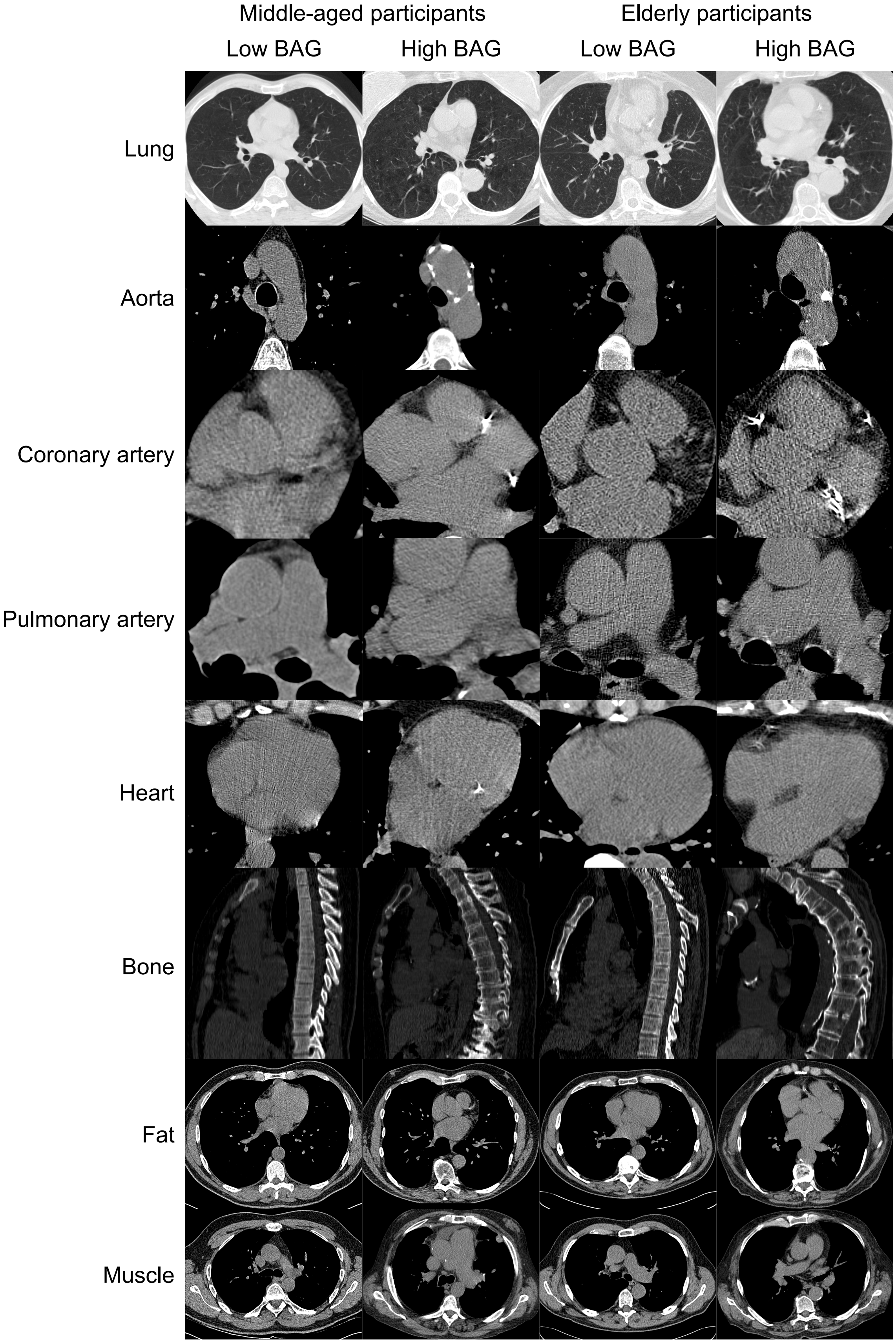

Middle-aged participants
Elderly participants
Low BAG
High BAG
Low BAG
High BAG
Lung
Aorta
Coronary artery
Pulmonary artery
Heart
Bone
Fat
Muscle


143

**Supplementary Fig. 1: Representative paired CT examples of low and high organ-specific BAGs at the same chronological age.**

Representative CT image crops are shown for eight thoracic organ and tissue compartments: lung, aorta, coronary artery, pulmonary artery, heart, bone, fat, and muscle. For each compartment, low- and high-BAG examples are shown as paired examples from participants with the same chronological age, separately within the middle-aged group aged 50–60 years and the elderly group aged 70–80 years. Within each same-age pair, low BAG indicates a younger-appearing CT phenotype, whereas high BAG indicates an older-appearing CT phenotype. BAG, biological age gap.

**Supplementary Table 1. Demographic and CT acquisition characteristics of the CT-RATE and MIDRC datasets used for model development.**

| **Cohort** | **CT-RATE** | **MIDRC** |
|---|---|---|
| Cohort size | | |
| Unique individuals, n | 2,829 | 4,083 |
| CT examinations, n | 5,494 | 4,477 |
| Demographic characteristics | | |
| Age used for model training, mean (SD), years | 33.1 (9.7) | 60.1 (16.9) |
| Female sex, n (%) | 2,534 (46.1%) | 2,253 (50.3%) |
| CT acquisition characteristics | | |
| Low-dose CT, n (%) | 249 (4.5%) | 96 (2.1%) |
| Contrast-enhanced CT, n (%) | 8 (0.1%) | 2,803 (62.6%) |
| Z-spacing, median (IQR), mm | 1.50 (0.75–1.50) | 0.76 (0.62–1.25) |

Data are mean (SD), median (IQR), or n (%). All characteristics were summarized at the CT examination level. Low-dose CT was defined as CTDIvol $<3.0$ mGy. Contrast-enhanced CT was defined by intravascular attenuation $\geq 80$ HU. HU, Hounsfield units; IQR, interquartile range; SD, standard deviation.

**Supplementary Table 2. Dataset availability of baseline comorbidities.**

| Baseline comorbidity | Dataset availability |
|---|---|
| High blood pressure | NLST and COPDGene |
| Diabetes | NLST and COPDGene |
| Stroke | NLST and COPDGene |
| COPD | NLST and COPDGene |
| Emphysema | NLST and COPDGene |
| Chronic bronchitis | NLST and COPDGene |
| Asthma | NLST and COPDGene |
| Breast cancer | NLST and COPDGene |
| Heart disease | NLST and COPDGene |
| Pneumonia | NLST only |
| Asbestosis | NLST only |
| Childhood asthma | NLST only |
| Bronchiectasis | NLST only |
| Tuberculosis | NLST only |
| High cholesterol | COPDGene only |
| TIA | COPDGene only |
| Sleep apnea | COPDGene only |
| Pneumothorax | COPDGene only |
| Compression fracture | COPDGene only |
| Interstitial lung abnormality | COPDGene only |

**Supplementary Table 3. Dataset availability of incident chronic diseases.**

| Incident disease outcome | Dataset availability |
|---|---|
| Lung cancer* | NLST only |
| COPD | NLST and COPDGene |
| Emphysema | NLST and COPDGene |
| Chronic bronchitis | NLST and COPDGene |
| Asthma | NLST and COPDGene |
| Sleep apnea | NLST and COPDGene |
| Atrial fibrillation | NLST and COPDGene |
| Stroke | NLST and COPDGene |
| Coronary artery disease | NLST and COPDGene |
| Heart attack | NLST and COPDGene |
| Congestive heart failure | NLST and COPDGene |
| Diabetes | NLST and COPDGene |
| Pulmonary fibrosis | NLST only |
| Pneumothorax | COPDGene only |
| Breast cancer | COPDGene only |
| Transient ischemic attack | COPDGene only |
| High blood pressure | COPDGene only |
| Osteoporosis | COPDGene only |
| Compression fracture | COPDGene only |
| Rheumatoid arthritis | COPDGene only |
| High cholesterol | COPDGene only |

*Lung cancer was analyzed using Cox proportional hazards regression because time-to-event data were available in NLST; all other incident disease outcomes were analyzed using logistic regression.

**Supplementary Table 4. ICD code-based classification of cause-specific mortality.**

| Cause-specific mortality | ICD codes |
|---|---|
| Respiratory | J* |
| Cardiovascular | I* |
| Cancer | C*; D0-D4* |
| Other | A047; A099; A310; A410; A412; A415; A419; A430; A481; A499; A810; B171; B182; B201; B203; B208; B212; B222; B227; B24X; B259; B344; B348; B379; D589; D609; D649; D65; D683; D691; D70; D735; D739; D801; D869; E039; E105; E109; E112; E115; E117; E119; E141; E142; E144; E145; E146; E147; E148; E149; E162; E232; E271; E43; E45; E46; E668; E669; E780; E785; E835; E854; E859; E86X; E872; E875; E880; E889; F011; F019; F03; F03X; F069; F101; F102; F104; F151; F172; F179; F191; F199; G001; G009; G039; G062; G10; G122; G20; G20X; G210; G309; G310; G311; G318; G319; G35; G35X; G409; G419; G473; G610; G700; G710; G728; G825; G903; G931; G934; G935; G939; G959; H709; K088; K222; K223; K228; K254; K255; K264; K274; K275; K279; K296; K319; K460; K519; K550; K559; K566; K572; K578; K579; K631; K635; K650; K659; K700; K703; K704; K709; K729; K746; K759; K768; K769; K800; K802; K810; K829; K830; K859; K860; K868; K900; K922; L031; L039; L120; L899; L89X; L930; M051; M069; M317; M329; M348; M349; M478; M503; M513; M608; M869; N049; N059; N179; N180; N185; N189; N19; N19X; N288; N289; N390; N839; Q282; Q613; Q822; R02X; R068; R092; R198; R54; R570; R578; R579; R580; R58X; R628; R688; R91; R960; R97X; R99; R99X; T179; V031; V039; V092; V239; V244; V270; V274; V294; V435; V436; V445; V470; V475; V481; V485; V486; V490; V494; V496; V499; V575; V58X; V689; V869; V877; V892; V959; V970; W08X; W10; W100; W10X; W11X; W170; |

| | |
|---|---|
| | W18; W18X; W19; W190; W199; W19X; W20; W20X; W49X; W69; W74; W749; W79; W80; X00; X008; X00X; X09; X090; X31; X37; X41; X419; X42; X42X; X44; X45; X47X; X590; X599; X62; X64; X64X; X67X; X700; X70X; X72; X72X; X73; X73X; X74; X740; X748; X749; X74X; X84X; X910; X95X; Y09; Y120; Y14X; Y19X; Y24X; Y26; Y34; Y427; Y450; Y830; Y831; Y838; Y848; Y850; Y86 |

Prespecified ICD codes used to classify underlying causes of death into cause-specific mortality categories for the NLST mortality analyses.
